## Supplementary Materials for "The multi-dimensional challenges of controlling respiratory virus transmission in indoor spaces: Insights from the linkage of a microscopic pedestrian simulation and SARS-CoV-2 transmission model"

**SUPPORTING INFORMATION**

**S1 Text. Supporting Information**
**S1.1. Background information on indoor movement and transmission models**
**S1.2. Experiment setting for static contacts**
**S1.3. Description of SamenSlimOpen tool**
**S1.4. Case study description**
**S1.5. Parameter description in QVE-MOD**
**References for supporting information**
**S1 Fig. Screenshots of the SamenSlimOpen tool**
**S2 Fig. The case study restaurant layout**
**S3 Fig. The snapshot contamination map in the case study**
**S4 Fig. The contamination maps in the case study for ventilation and face mask scenarios**
**S5 Fig. The analysis of relative contribution of transmission routes in the case study**
**S6 Fig. Sensitivity analysis of emission rate**
**S7 Fig. Sensitivity analysis of proportions of aerosols**
**S8 Fig. Sensitivity analysis of virus decay rate on surfaces**
**S9 Fig. Sensitivity analysis of virus transfer rate between hand and surface**
**S10 Fig. Sensitivity analysis of fractional virus transfer rate from hand to facial membranes**
**S11 Fig. Sensitivity analysis of diffusion rate and deposition rate**
**S12 Fig. Sensitivity analysis of diffusion rate and virus decay rate in aerosols**

### S1 Text. Supporting Information

#### S1.1. Background information on indoor movement and transmission models

The following section describes the existing models. The first section (S1.1.1) discusses the state-of-the-art pertaining pedestrian strategic and tactical choice models for indoor environments. Section S1.1.2 continues with an elaboration on simulation models for operational movement behaviour. Section S1.1.3 reviews models that simulate the spread of respiratory viruses in indoor spaces. This section ends with an overview of the attempts to use pedestrian simulation models and virus spread models to determine SARS-CoV-2 transmission risks (Section S1.1.4).

##### S1.1.1. Modelling pedestrian strategic and tactical choice behaviours

The movements of individuals are governed by choices at three levels, namely strategic (activity, activity location, departure time choice), tactical (mode and route choice) and operational choices. In particular, this section will focus on the state-of-the-art of activity and route choice behaviour literature because these two choices are influential in the case of individuals' movement behaviour in indoor spaces. More specifically, we will focus on models that determine the order of individuals' activities and their routing.

###### *S1.1.1.1. Activity scheduling and location choice modelling*

Most studies focussed on activity choice modelling to represent the movements of pedestrians across city-scale networks. Borgers and Timmermans (2010) were one of the first to develop a set of three connected models that jointly identify pedestrian movements in shopping areas. Dijkstra and Jessurum (2016) simulate visitor's activity scheduling in shopping areas. Clifton et al. (2016) and Oyama and Hato (2018) determine pedestrian destination choices in a city-network. Vukadinovic et al. (2011) developed an activity simulation model for another leisure activity, namely theme parks. Danalet et al. (2014) and Danelet and Bierlaire (2014) adopted discrete choice models to identify the restaurant choices and activity patterns of students and staff of the university.

More recently, several studies featured activity choice modelling inside buildings. Hoogendoorn and Bovy (2004), for instance, developed a continuous utility maximisation framework that makes a trade-off between activity performance and the waiting and walking costs. This model is often applied in real-life cases, where the parameters of the activity utility were estimated using data of a benchmark situation. Ton et al. (2015) established a set of single- and multi-factor MNL models to identify the activity schedules of travellers inside train stations. Kalakou et al. (2014) applies the same approach to model activity schedules at airports. Liu et al. (2014) used a nested logit model to predict activity travel patterns for airport travellers. Shelat et al. (2020) developed a markov-chain model to simulate the activity scheduling of individuals working in an office building. The transition probabilities were derived from movement data inside the building.

What all of these activity location & scheduling modelling approaches have in common is that they require large amounts of data to be correctly estimated and that they are often difficult to generalise. Yet, for indoor scenarios, and specifically restaurants, little data is available regarding individuals' activity scheduling. Thus, the modelling approaches presented above are difficult to use. Yet, the literature also shows that if one can estimate the transition probabilities between activities, the resulting activity scheduling behaviour is relatively realistic. Thus, in this first, pragmatic attempt to model activity choices in restaurants, an expert-driven approach is adopted. For more information, see section 5.1.

##### *S1.1.1.2 Route choice modelling*

Early works on pedestrian route choice modelling, such as Hill (1984), indicate that route choice selection strategies are largely subconscious, but that directness is the most commonly identified reason for choosing a particular route. Various studies have afterwards identified the impact of the built environment on pedestrian route choices in city-scale networks (e.g. Borgers and Timmermans (2005), Guo et al. (2013), Lue and Miller (2019), Shatu et al. (2019)). Often discrete choice models, such as the path size logic model and the multinomial probit models, are estimated. Some, more data-driven approaches directly infer the likelihood of a given path based on a large set of realised traces and their characteristics, such as speed distribution and nearby WiFi spots (e.g. Chen and Bierlaire (2015)).

Yet, with respect to indoor route choice behaviour, most works empirically studied indoor navigation, see Kalakou and Moura (2014) for an overview. Additionally, some utility-based route choice models have been developed for specific contexts (e.g. Cheung and Lam (1998), Hughes (2000), Hoogendoorn and Bovy (2005) and Daamen et al. (2005)). Yet, given that the models are difficult to generalise and/or estimate due to their high data requirement. Consequently, the shortest route towards one's destination is frequently adopted in simulation approaches. Often local biases or forces are introduced to account for crowding at the scene (e.g., Campanella et al. (2009), Usher et al. (2010)). The adoption of the shortest-distance path can, however, lead to unrealistic 'suboptimal' pedestrian choice behaviour. Asano et al. (2010), Kretz et al. (2011) and Stubenschrott et al. (2014) developed more sophisticated route choice models that determine the shortest path dynamically, thus improving local route choice.

Even though dynamic route choice models are most sophisticated, their computational effort is relatively high. Moreover, in case of low demand locations (due to physical distancing restrictions), we expect the adverse effects of suboptimal routing to be limited. Thus, because we favour fast computation times and expect limited routing issues, this study will apply the utility-based routing approach presented by Hoogendoorn and Bovy (2004), also coined Nomad, to globally route pedestrians between activity locations.

#### S1.1.2. Modelling individuals' operational movement dynamics

Over the years various pedestrian movement models have been presented. The level of details pertaining to the spatial representation of the environment, agents' choice behaviour and agents' movement behaviour differs greatly. Six distinctive types of models can be identified, namely i) Graph-based models, ii) Continuous models, iii) Cellular Automata models, iv) Force-base models, v) Velocity-based models and vi) discrete choice models. All model types are discussed below, for a more in-depth discussion of the (dis)advantages of all model types the reader is referred to Duives et al. (2013).

The first two types both make use of a macroscopic representation of pedestrian flows, using the fundamental relation between density and walking speed (P-FD) to determine the flow of pedestrians across boundaries. *Graph-based models* simulate the movements of pedestrians along a simplified graph (e.g. Chalmet et al. (1982), Hanseler et al. (2017)) consisting of vertices and nodes. *Continuous pedestrian simulation models* discretize pedestrian spaces in cells along 2 dimensions, but still make use of a continuous representation of pedestrian flows (Hughes (2000; 2002), Treuille et al. (2006)). Few macroscopic models can cope with heterogeneous crowds and multi-directional traffic, amongst which Hoogendoorn et al. (2014), Tordeux et al. (2018)). Continuous macroscopic models are best used for scenarios where large crowds move through relatively confined simply-shaped spaces.

Besides that, four major streams of microscopic pedestrian simulation models have been developed in the last four decades. These models all simulate the movements of discrete pedestrians. *Cellular automata models* were first developed by Blue and Adler (1998), and have been further specialized ever since (e.g. Song et al. (2005), Schadschneider (2002), Sarmady (2010), Bandini et al (2011)). The environment is often spatially discretized in a regular grid, which can limit CA's validity in high-density situations and small or oddly shaped spaces. *Force-based models* have been on the rise since the seminal paper of Helbing et al. (1995). This second model type determines the next position of a pedestrian as the result of its current movement direction and force-based interactions with other agents, its destination and its surrounding. This popular model type has seen many extensions, amongst which other force-interpretations (e.g. Hoogendoorn and Bovy (2004)), interaction biases (e.g. Parisi et al. (2009), Chen et al. (2020)), stairway behaviour (e.g. Qu et al. (2014)), corner rounding behaviour (e.g. Chen et al. (2020), Dias et al. (2019)), travel time (Kretz et al. (2011)). *Velocity-based models* determine the next speed of an agent as the result of the agent's destination, kinetic energy dispensal, and collision-risk (Karamouzas and Overmars (2010), Mousssaid et al. (2011), Paris et al. (2007)). Interactions directly influence the velocity of agent's, which tends to make agents' movements fairly unstable. The last type of microscopic model, i.e. *discrete choice models*, simulate pedestrians' operational movements as an optimization of its choice utility (e.g. Antonini et al. (2004), Robin et al. (2009)), which are the weighted sum of various elements of the potential next positions. This model type is not often applied, as it is very difficult to create a flexible 'simple' movement model that can cope with the general interaction complexity of public spaces.

For the modelling of virus transmission, a detailed description of pedestrians' movements is essential. Thus, a microscopic model is better suited to simulate the necessary movements. Additionally, most SARS-CoV-2 transmissions occur in indoor spaces, where a multitude of complex interaction behaviours occur. This vouches for a microscopic model for which many specialist behaviours have already been implemented (e.g. force-base or velocity-based). At the same time, the current physical distancing rules require the model to correctly simulate complex social interactions at a distance (no physical forces). A force-based model is better equipped to handle social interactions at a distance due to its setup. Thus, a generalisation of a force-base model will be used to simulate the pedestrian choice and movement dynamics in indoor spaces, in particular Nomad (Hoogendoorn and Bovy (2004)) in the implementation of Campanella et al. (2014).

**S1.1.3. Modelling indoor transmission of respiratory viruses**

Indoor transmission models have been developed to better understand transmission of respiratory viruses and assess the effectiveness of non-pharmaceutical interventions (NPIs) (Kriegel et al. 2021; Gao et al. 2021). Models differ in the types and extent of heterogeneity that is taken into account. In general, three main aspects are considered, namely i) the virus distribution in the environment, which can be assumed well-mixed or distributed in space according to underlying biophysical mechanisms, and individual homogeneity or heterogeneity in terms of ii) infectiousness and/or susceptibility, and iii) in terms of human movement. What model is best suited, depends on the objectives of the research. In this section, we discuss the pros and cons of each model type.

In the most simple family of airborne transmission models, the Well-Riley models, one assumes that infectious particles are well-mixed and the quantities are at steady-state. It uses an analytical expression, the Well-Riley equation, to relate the number of infected and susceptible individuals in an indoor space, the ventilation rate and the amount of infectious particles in the air to the expected number of new cases over a certain time period. Wells (1955) used a hypothetical infectious dose unit "quantum of infection" to describe how many infectious particles an infected individual emits. A quantum unit is defined as the number of infectious particles required to cause 63% of people getting infected, and is usually back- calculated using data on numbers of individuals infected in well defined indoor outbreaks. The Well-Riley model has been extensively used for predicting the number of infected individuals expected at large gatherings and assessing the effect of interventions such as capping the occupancy and reducing the total event duration (Bazant and Bush 2021; Kriegel et al. 2021). It is often used for quick risk assessments of large events. However, since every individual has the same risk of infection, the Well-Riley models do not allow for assessing the role of individual behaviours (contact frequency and respiratory activities e.g.,) on transmission risks.

Aerosol infectious dose-response models allow individual heterogeneity in infectiousness and respiratory activities. These models model the risk of getting infected from being exposed to different amounts of an airborne pathogen. This exposure dose, the amount of pathogen that reaches the susceptible individuals, depends on the infectious people's emission rates, the room volume, the recipients' pulmonary ventilation rate (i.e., the amount of air inhaled per unit time), and the exposure duration. Like the Well-Riley model, these models assume infectious particles to be distributed homogeneously with the virions concentration equal to the ratio between the emitted infectious particles and the room volume. The probability of infection occurring given exposure to a certain dose can be modelled using mathematical formulations, e.g., exponential dose-response relationship (Sze To and Chao 2010). This approach is more flexible than Well-Riley models since it allows for individual-level heterogeneity in terms of infectiousness, respiratory activities, and exposure time, and can be used to calculate individuals' infection risk (Lelieveld et al. 2020). This approach can be used to compare the relative exposure risk of common indoor environments such as classroom and office (Jones et al. 2021). However, the assumed homogeneous spatial distribution of infectious particles has the consequence that the distance between susceptible and infectious individuals does not affect exposure to the virus. Therefore, as with the Well-Riley models, the impact of physical distancing cannot be evaluated with this family of models.

Recently, more detailed infectious dose models have been developed that allow for spatial variation of the pathogen's distribution in the environment as well as multiple transmission routes (Gao et al. 2021). Expansions of the Well-Riley model have been proposed that allow for individual heterogeneity in infectiousness and respiratory activities (Bazant and Bush 2021; C. Xu et al. 2022), for spatial variation of the virus distribution in the environment (Lau et al. 2022; X. Li et al. 2022), and the inclusion of multiple transmission routes (Gao et al. 2021). In addition to airborne transmission, which was implicitly assumed in models mentioned above, these models consider transmission via droplets (i.e., larger respiratory particles that do not remain airborne) and fomites (i.e. contaminated objects). The inclusion of different transmission routes allows for increased granularity of the spatial and temporal dynamics of respiratory disease pathogens in their environments. In an early example of this model family, Nicas and Sun model virus-laden aerosols to be evenly distributed in a room, while viruses in droplets only distribute near the infectious individual in their respiratory cone puff (Nicas and Sun 2006). The distribution of virus on fomites in turn depends on the deposition of these viral-laden droplets and may accumulate in time depending on the duration of stay of the infectious individual. This framework is formulated by discrete events and can be expanded on the details of the various processes involved, such as pathogens emission, pathogens survival in the environment, recipients' uptake and causation of infection (Atkinson and Wein 2008; Sheng Li et al. 2009; Spicknall et al. 2010). Several event-based multiple routes infectious dose models have been built for COVID-19 (Arav, Klausner, and Fattal 2020; Gao et al. 2021), which shed light on how dominant routes depend on the duration and distance of infectious contacts.

For the highly micro level but computationally-intense modelling of virus-laden airborne particles, one group of indoor transmission models follow the computational fluid dynamics (CFD) principles and simulate the flow of particles indoors in time and space (Mirzaie et al. 2021; H. Liu et al. 2021; Ren et al. 2021). These models have a clear advantage in detailed modelling of the distribution of particles, and allow for additional complexity such as airflows due to open windows. However, the computational requirements of these models limit their use for larger and longer lasting events.

##### S1.1.4. Simultaneous modelling of individuals movement behaviour and SARS-CoV-2 transmission

The models summarised typically consider static contacts between infected and susceptible individuals and thereby ignore the dynamic nature of human interactions. Combining detailed pedestrian models with viral transmission models can counter this issue and help investigate the role of the contact networks in indoor spaces.

Pedestrian movement models have been integrated with viral transmission models before. Sallah et al. (2017) built a mathematical model that combines a graph-based mobility model and a SIR model to predict the spread of cholera in Haiti. While this model simulated the spread on (sub)national levels, other models are focussed on small-scale transmission, such as in indoor spaces. Harweg et al. (2021), Yang et al. (2020), Xiao et al. (2021), Romero et al. (2020), Ronchi et al. (2020), Xu and Chraibi (2020) developed hybrid models that combine microscopic movement models for simulating pedestrian dynamics with viral transmission models. Most often, the distance and duration of, what is considered an infectious contact, is predefined. One group of hybrid models estimate the exposure time, as it relates to different definitions of 'risky contacts'. In the case of Xiao et al. (2021) a force-based model was built to model pedestrian movement, and the total duration of risky contacts was calculated to assess the impact of interventions such as shortening opening hours. Similarly, in Fang et al. (2020), the duration of risky contacts is calculated based on cut-off distance, with different cut-off distances assumed for breathing and coughing (one metre and two and a half metres, respectively). Ronchi et al. (2020) classified exposure into five types with different distances, which reflect different transmission mechanisms. The exposure time for each type of exposure was calculated to show the general exposure risk in the same indoor space.

Another group of hybrid models estimates infection probabilities, either by defining a probability per risky contact or assuming infection probabilities to be linearly related to the exposure time. Fang et al. (2020) applied an infection probability curve that increases with the duration of the contact and assumed that people can get infected only when they have close contact with infectious individuals (less than one metre), while Bouchnita and Jebrane (2020) assumed a cut-off distance of two metres. The total exposure time is calculated based on cut-off distance and compared to a confirmed duration within which close contact will definitely cause infection. Similarly, Ying and O'Clery (2021) model the infection probability of an exposed customer as a linear function of the exposure time, depending on the duration that the susceptible and the infectious agents spend in the same zone. The relation between exposure and infection is, however, hard to calibrate against epidemiological data and often not done explicitly.

Most hybrid models presented above predominantly assume contact transmission and ignore indirect transmission routes such as via aerosols and fomites. We will add to the state-of-the-art by developing an individual-based model that combines a pedestrian movement model and a multi-route transmission model. By allowing for spatial and temporal heterogeneity in both human behaviour and viral spread, we aim to better understand the impact of human behaviours on transmission risks.

**S1.2. Experiment setting for static contacts**

In the static contact experiments (Fig 2), susceptible and infectious individuals stand still in a 10*10 m indoor space. There are tables next to both individuals, but individuals do not share any common surfaces. The benchmark contact is defined as a scenario where susceptible and infectious individuals arrive concurrently in an indoor space and have a contact at a distance of 1.5 metres for 15 minutes, which is broadly used as an indicator of 'a risky contact' (RIVM 2021). Both infectious and susceptible individuals are assumed to talk and breathe for 50% of the time each (akin to an interaction in a restaurant for instance). The indoor space has an average ventilation level of 3 ACH (air change per hour).

To examine the impact of contact intensity on exposure, we conduct three experiments in Section 3.1.1. The first experiment compares exposure at different contact distances with the benchmark contact settings. The susceptible individual is placed at different distances away from the infectious individuals, from 0.5 metres to 4 metres, to examine the impact of contact distances. In the second experiment, we examine the impact of contact duration by placing susceptible and infectious individuals in the indoor space for different durations from 5 mins to 120 mins, while the rest of the settings are the same as the benchmark contact settings. Finally, we repeat the first two experiments but introduce the infectious individual in the indoor space 3 hours prior to the susceptible individuals to examine the impact of virus contamination accumulation in the environment on exposure.

We thereafter explored the impact of respiratory activities on transmission between static contacts. Compared to breathing, active respiratory activities, such as singing and talking loudly, are associated with increased emission and subsequent exposure to viral particles by proximate susceptible individuals (Hamner et al. 2020; Charlotte 2020). Moreover, active respiratory activities result in higher tidal volume (inhalation rate), which can further increase virus exposure. We used estimates from exhaled breath samples by (Coleman, Tay, Sen Tan, et al. 2021) and a systematic literature review (P. Z. Chen et al. 2021) to parametrize emission, particle composition, and inhalation upon different respiratory activities (See Table 3). We simulated exposure upon the benchmark contact (1.5 metres for 15 minutes) as a result of 15 minutes of the respective activity. Specifically, in the experiments in Section 3.1.2, the susceptible and infectious individuals are placed in an indoor space with benchmark contact settings, and both the infectious and susceptible individuals only conduct one type of respiratory activity for 15 minutes (either breathing, talking or singing).

Interventions such as ventilation and facemasks are also implemented in the PeDVis model. Susceptible and infectious individuals are placed in an indoor space with a benchmark contact set-up under different ventilation settings. Ventilation was modelled assuming a constant, spatially homogeneous renewal of air. This is reflected by the air change rate per hour (ACH), which is a measure of the number of times the air in a space is fully replaced per hour. For reference, the ACH in Dutch residential dwellings is roughly 0.1-1.9 (Van Ginkel and Hasselaar 2006) versus 3 in most indoor public settings such as classrooms and supermarkets (CIRES 2020). The recommended ACH in home and business is 6 (Arboportaal 2020; RIVM 2020). We assumed that ventilation only affects the amount of viral-laden aerosols built up in the environment. In the first set of experiments in Section 3.1.3, susceptible and infectious individuals are placed in a benchmark contact setting with half-time breathing and half-time talking. The ventilation of the room is set to changing values from 0 to 25.

Lastly, in the second set of experiments in section 3.1.3, we examine the impact of different facemasks on exposure in a benchmark contact. Face masks can influence both the emission and inhalation processes. The reduction of emission and inhalation rate is determined by face masks' filter efficiency (FE, the proportion of particles filtered out) (Bezek et al. 2021; Drewnick et al. 2021). This depends on the materials used, among others, and can range from as low as 20% for thin acrylic mask to more than 95% for N95 masks (Pan et al. 2020). Mask fit is also an important indicator of FE (Darby et al. 2021). As face masks reduce the number of big particles more than small particles emitted by breathing and talking (Asadi et al. 2020), we assume that face masks reduce the large particles ten times more efficiently than they do small particles (i.e., $(1-FE_{droplets}) = 10*(1-FE_{aerosols})$) (Asadi et al. 2020). Unless otherwise stated, we used the filter efficiency values given in Table 3 in the experiments. Additionally, we simulated a benchmark contact with both infectious and susceptible individuals wearing a mask with FE ranging from 0% to 100% for aerosols and the corresponding FE for the droplets.

**S1.3. Description of SamenSlimOpen tool**

The PeDViS model is also available as an online tool (see SamenSlimOpen.nl/de-tool for the latest release), specialised for impact assessment of restaurants (see S1 Fig). S1 Fig presents a visualisation of the web-based interface via which users can design and simulate a restaurant. Users are asked to make a simplified representation of their venue in an accessible manner. The users can select a scenario or make one themselves which determines the length of the simulation and the occupancy over time, among others. After setting up their restaurant, the users start the simulation. The output of the simulation is a heatmap of the cumulative virus exposures that occurred over the course of the simulation, as well as summary statistics of the simulation.

The goal of the SamenSlimOpen project is not only to inform users about the relative virus exposure risks of their current setup but also to learn about what measures to use in conjunction to limit SARS-CoV-2 transmission in their venue. The SSO app allows users to try different configurations and compare results. Users can change their restaurant layout or change the scenario: the occupancy, how long guests are staying, or what measures are taken.

**Table A. Implemented measures in the SSO app to limit SARS-CoV-2 transmission**

| Measure | Option A | Option B |
|---|---|---|
| Face masks | Guests don't wear masks | Guests wear masks when they walk |
| Payment | At the register | At the table |
| Coats | On the coat rack | Taken to the table |
| Shifts | Tables are used multiple times per evening | Tables are used once per evening |
| Air change rate | A continuous slider which sets the times of air change per hour | |
| Cleaning tables | A continuous slider which sets the interval time of tables being cleaned | |

In order for the SSO app to simulate, the user input for the PeDVis model is collected in the graphical user interface. The user first identifies the layout of the restaurant. Here, the location of all objects and functional spaces (e.g., toilet, coat rack, cash register) are determined. All objects can be scaled and rotated to best resemble the layout of their restaurant. The number of tables and chairs placed in the restaurant constrain the number and sizes of groups that can be accommodated in the restaurant. Moreover, their placement and size affect the walking routes of the simulated individuals. Accordingly, the user indicates the actual maximum occupancy, the average duration of stay in the restaurant, and the time slots at which groups are expected to arrive at the restaurant. Lastly, the user can indicate whether to use intervention measures, including wearing face masks when walking (Table A).

**S1.4. Case study description**
In the presented case study, the SSO app was used to create the restaurant scenario's. The underlying PeDViS model was then used to simulate and compare the impact of various measures to limit SARS-CoV-2 spread in restaurants.

For this case study, a small (10 * 9 meters) conceptual restaurant is adopted (S2 Fig). The restaurant contains four tables, which are distributed over the space. Each table has four chairs, all of which can be simultaneously occupied by visitors. The restaurant also has a bar with four stools. The capacity is determined by the number of chairs/stools in the restaurant, so twenty visitors can be present simultaneously . The main entrance and exit of the restaurant are on the northern side of the restaurant, with a coat rack next to the entrance. The cash register is on the left side of the bar, and two toilets are located on the southern side of the restaurant. The material of the chairs, tables, and bar is assumed to be wood. The objects in the room are disinfected one time before the restaurant opens, and not cleaned again during the following six-hour shift.

At the restaurant, the main activity of all individuals is sitting on a chair. Besides that, individuals can perform three intermediate activities. They can hang their coat on the coat rack, go to the toilet, or pay at the cash register. In the case study below, all individuals hang their coats on chairs and pay at tables. The probability that an individual visits the toilet is assumed to be 0.6. The timing to go to the toilet is random, and the time spent there is drawn from a normal gaussian distribution with a mean of 120 seconds and a standard deviation of 60 seconds, with a maximum of 240 seconds. Consequently, individuals move to these two locations and thereafter move back to their main activity location.

The simulated walking behaviour of the population is homogeneous in all aspects, except for the desired walking speed. In particular, the same parameter settings are used for each individual in the crowd. As a result, all agents have a similar mass, understanding of the surrounding space, collision avoidance, rotation, and acceleration behaviour. For each individual, the desired walking speed is drawn from a normal gaussian distribution with a mean speed of 0.9 m/s and a standard deviation of 0.2 m/s. The maximum speed is set to 1.4 m/s. They are not obliged to comply with the 1.5 metre distancing rules, which means individuals will use all available space to move. This is in line with observations from crowd monitoring that individuals in confined indoor spaces have difficulty complying to distancing rules and no big differences in pedestrian behaviour despite the introduction of distancing measures (van Schaik et al. 2022; Villena Gonzales 2022).

The simulation lasts for 6 hours of service at a restaurant, in which some tables are used twice. Thus, in one complete simulation run, 32 individuals have entered the simulation. At the end of the simulation, some individuals might still be present in the restaurant.

Only one infectious individual enters the simulation during its runtime, which is randomly assigned at the beginning of QVEmod. For that reason, the SSO app normally runs in multiple replications and then generates results for the expected number of infections. However, for the case study results to be comparable, being infectious is assigned to a fixed agent (ID= 9), the time spent by whom at the restaurant corresponds to approximately the middle part of the simulation runtime (enters the restaurant approximately at the beginning of the 2nd hour and leaves at the beginning of the 4th hour) (Fig 5C), and sits on a table which is approximately in the middle of the restaurant (Fig 5A). All other characteristics are similar to that of other individuals and are randomly drawn at the entrance of the simulation.

**S1.5. Parameter description in QVE-MOD**

**Emission rate** ($\omega$) represents the relative viral load of an infector emitting per hour. In the case study, $\omega$ is scaled to 1 per hour for a typical infectious person with a typical respiratory activity for the restaurant context, which is assumed to be half-time breathing and half-time talking.

**Emission quantity** ($\phi$) is set to a default value of $10^6$ copies per hour. This value is based on work done by Ma et al. (2020). They collected 20 minutes of exhaled breath samples from COVID-19 patients and estimated emissions to range from $1.13*10^5$ to $2.25*10^7$ copies per hour (Ma et al. 2020). This range was in agreement with other studies including that of (Evans 2020), who measured viral load in the saliva (1000 /nl) and multiplied this with the average volume of emitted saliva per hour (330 nl/hour). This resulted in estimates of around $3.3*10^5$ copies per hour, when people spend half of their time talking and the other half breathing (Evans 2020). A more recent study quantified that the emission rate from omicron patients is around $5*10^7$ copies per hour (Zheng et al. 2022). As there is substantial heterogeneity in emission quantity between individuals, we performed a sensitivity analysis spanning emission rates for $10^5$ to $10^7$ copies (S6 Fig).

**Respiratory activity scaler** ($\delta$) is a relative scale representing the differences in emission rates among different respiratory activities. It is quantified based on work by Coleman et al. (2021) who specified the relative difference of RNA copies in breath samples (n=13) during the different respiratory activities. According to the study, an individual singing emits 16415.5 RNA copies during 15 minutes of continuous singing, while emitting 12655.9 RNA copies during 15 minutes of talking and 1959.3 copies during 30 minutes of tidal breathing. These findings indicate that singing and talking result in 16.7 times and 12.9 times more viral particle emissions, respectively, compared to breathing. In our simulation experiments, the default respiratory activity is defined as half-time breathing and half-time talking and the corresponding respiratory activity scaler ($\delta$) is set to 1. The respiratory activity scaler for other scenarios can be calculated based on the emission rates of different activities relative to the default activity. We used the findings from Coleman, Tay and Tan et al. (2021) in our simulation experiments to calculate these relative activity scalers. Other studies that shed light on the relative emission by respiratory activity include (Gregson et al. 2021; Mürbe et al. 2021; Schijven et al. 2021). These measured the particle mass, but not the virus amount emitted, during different respiratory activities and were considered less direct estimates for this parameter.

**The infectiousness scaler** ($\sigma$) is used to scale individual infectiousness. 0 represents a susceptible individual who cannot emit the virus and 1 represents a “typical infectious” individual. Values of $\sigma$ that are greatly larger than 1 can be used to reflect super-shedding individuals.

**The proportions of aerosols ($p_{aerosols}$) and droplets ($p_{droplets}$) during emission** represent how the emitted viral particles are partitioned in aerosols and droplets, respectively. These were parameterized based on three factors: i) a predetermined cut-off size between aerosols and droplets, ii) the relative amounts of aerosols and droplets expelled by humans, and iii) the difference in viral copies carried by aerosol and droplet particles (P. Z. Chen et al. 2021). According to the results presented in the systematic literature review studies (Pöhlker et al. 2021; P. Z. Chen et al. 2021), particles can be classified into droplets (>100um), buoyant aerosols (<10um), short-range aerosols (50um<d<100um) and long-range aerosols (10um<d<50um). Recent studies that used laser particle counters to quantify the respiratory particles’ size distribution among different respiratory activities showed that most of the particles could be classified as aerosols having small sizes (Mürbe et al. 2021). In our study, we defined one cut-off size for the particles, and classified them into two categories: i) buoyant aerosol particles having a dry size of < 10um (assumed to float in the air layer), and ii) droplets having a dry size of > 10um (assumed to float in the air first but then fall onto the surface layer in time). The calculated proportions in different respiratory activities and the corresponding respiratory activity scaler values are listed in Table B. When different scenarios require changing the composition of respiratory activities, the proportions can be recalculated by considering the duration-based weights of each activity and the corresponding respiratory activity scaler. For example, $p_{aerosol}$ in a scenario including a different set of respiratory activities can be calculated as shown below.

$$p_{aerosol,scenario(activity)} = \frac{\sum p_{aerosol,activity} * \delta_{activity} * duration_{activity}}{\sum \delta_{activity} * duration_{activity}}$$

For sensitivity analysis purposes, we tested the impact of different proportions of aerosols and droplets on our case study results and reperformed the case study simulations with changing aerosol proportions of 50% lower and higher values with corresponding droplet proportions (S7 Fig).

**Table B. The proportions of particles in aerosols and droplets among different respiratory activities**

| Scenario | p (aerosols) | p (droplets) | Respiratory activity scaler |
|---|---|---|---|
| Talking | 17.1% | 82.9% | 1.86 |
| Singing | 6.52% | 93.48% | 2.4 |
| Breathing | 97.8% | 2.2% | 0.14 |
| Half-time breathing + half-time talking | 22.9% | 77.1% | 1 |

**Unit decay rates of virus on surfaces ($\mu_{fomites}$) and aerosols** ($\mu_{aerosols}$) were set based on experimental studies investigating the stability of SARS-CoV-2 in the environment. The survival of SARS-CoV-2 in different environmental conditions depends on the type of surface, the presence of organic matrix, temperature, and humidity (Chin et al. 2020; Ijaz et al. 2021; Kasloff et al. 2021; Pastorino et al. 2020; Matson et al. 2020; Harbourt et al. 2020; Y. Liu et al. 2021; Riddell et al. 2020; Ranjan 2022). The viral particles lose infectiousness at decay rates that depend on the environment and the surface material. The decay rates can be calculated based on experimental studies estimating the half-life of particles.

In our case study, the main material used for the surfaces is *wood*, with an exponential decay rate of **0.969** $hr^{-1}$ (Chin et al. 2020). The surface types in our model are configurable to common materials used in restaurant settings, such as plastic, glass, cloth, paper, cardboard, copper, and steel, and the corresponding decay rates are calculated using literature (van Doremalen et al. 2020; Y. Liu et al. 2021). Considering the range of decay rates on different surfaces, a sensitivity analysis was performed within the -90% and +90% range of the base decay rate (S8 Fig).

Estimates for decay rates of SARS-CoV-2 particles in aerosols vary greatly. According to van Doremalen et al. (2020), using TCID50 experiments, SARS-CoV-2 can survive in the suspended aerosols for a few hours with a median half-life of 1.09 hours which corresponds to an exponential decay rate of **0.636** $hr^{-1}$. In another study with TCID50 experiments, Smither et al. (2020) observed half-life durations between 0.50-1.25 hours (indicating decay rates between **0.555-1.386** $hr^{-1}$) using tissue culture media under different relative humidity conditions. Dabisch et al. (2021) found that in an environment without sunlight at 30 °C temperature and 70% relative humidity (which represents an approximate condition that can also be observed in indoor settings), the time needed for a 90% decrease in infectious virus is 35 minutes, which corresponds to a decay rate of **3.947** $hr^{-1}$. In a more recent study, Oswin et al. (2022) found that the airborne infectivity of SARS-CoV-2 falls to 50% within 5 seconds on average in medium humidity conditions and 5 minutes in high humidity conditions, and observed a decrease in infectivity to approximately 10% in 20 minutes in both cases, which corresponds to a decay rate as high as **6.908** $hr^{-1}$. Considering this range of estimates, we applied a default decay rate of 1.5 $hr^{-1}$, which corresponds to a half life of about half an hour. To assess the sensitivity to this assumption, we applied aerosol decay rates 50% lower and higher than the default to our case study (S9 Fig). Since the decay rate is directly related to how long aerosols remain airborne and thus how far they get to travel, we performed the sensitivity analysis of the aerosol decay rate across levels of the diffusion coefficient (S9 Fig).

**Infectious dose ($D_{inf}$)** is the minimum dose needed to reach the target cells to cause an infection. It is set to 1000 based on estimates of the founding virus population size required to cause infection in a recipient host (Popa et al. 2020).

$c_{routes}$ is the proportion of viral particles someone is exposed to that reach the respiratory tract cells by route (aerosols, droplets, fomites). The baseline c values are assumed to be 10% for all the transmission routes based on the model analysis results in presented Section 3.2.3 and given in Table 1. This assumption aligns with other modelling studies and studies on aerosol deposition in lungs (Zuo, Uspal, and Wei 2020; Hinds 1999; Kraay et al. 2021).

**The volume of a cell (L)** is the volume of a grid cell, with a default value of 125 L, since each side of grid cells has a default length of 0.5 m.

**Air change rate (ACH)** is a measure of how many times the air in a defined space is fully replaced per hour. In our study, it takes values between 0-24 depending on the simulation experiment settings. ACH is estimated to be between 0.5 to 1.5 inside houses and between 3 to 15 with windows open (CIRES 2020).

**Filter efficiency (FE)** represents the proportion of particles filtered out in case of mask use. In our model, FE is used to calculate the reduction of emission and inhalation rate. The baseline FE is assumed to be 40% for aerosols (Ueki et al. 2020) and 94% for droplets. Here, we assume that the FE of both particle types are related according to (1-$FE_{droplets}$) = 10%*(1-$FE_{aerosols}$).

**Unit deposition rate of viruses in droplets** ($\mu_{droplets}$) is the deposition rate of droplets from the air layer onto surfaces. This is a physical process where the sedimentation time depends on the height, and particle size (Xie et al. 2007). The sedimentation time is proportional to the particle size d: ($1/d^2$). In this study, particles with d>10 um are classified as droplets. The average sedimentation time of these particles is calculated by using the distribution of the particle sizes that vary across different respiratory activities. In order to estimate the average sedimentation time, the particle size distribution data were extracted from the distribution figures for different activities (Morawska et al. 2009). The corresponding sedimentation time for each particle size was paired and the expected average sedimentation times were calculated for each respiratory activity based on the probability distribution of particle sizes. Considering the particle size distribution during each activity, the average sedimentation time for breathing, talking, and singing activities are calculated as 88.22 s, 101.64 s, and 137.46 s, respectively. Then, the corresponding deposition rates are quantified as 40.8 per hour, 35.4 per hour, and 20.18 per hour. Since the baseline scenario represents half an hour of breathing and half an hour of talking, the deposition rate in the baseline scenario is calculated as 37.93 per hour or otherwise put, half of the particles have deposited after 0.8 minutes.

For sensitivity analysis purposes, we tested the impact of different deposition rates on our case study results and reperformed the case study simulations with deposition rates of 50% lower and higher values (S10 Fig). Since the deposition rate is directly related to the time spent in the air by droplets, the sensitivity analysis of deposition rates is performed with the combined effects of changing diffusion coefficient values (S10 Fig).

**Diffusion coefficient** ($D$) defines how fast viruses diffuse in the air. Both droplets and aerosols can disperse into their surroundings. For diffusion equations, we adapted a model from Vuorinen et al. (2020), who investigated aerosol transport in indoor spaces and built a Monte-Carlo model to incorporate spatial-temporal aerosol dispersion. The diffusion coefficient used in Vuorinen et al. (2020) is $D$ = 0.05 $m^2/s$, whereas an experimental study reports that the effective diffusion coefficient value for aerosol particles is 0.0016 $m^2/s$ (O. B. Kudryashova et al. 2015). A later study suggests that the effective diffusion coefficient for SARS-CoV-2 can be set to 0.0016 $m^2/s$, but can be high as 0.01 $m^2/s$ when the convection is high in the indoor space (Olga B. Kudryashova et al. 2021). We set *D* to 0.0016 $m^2/s$ as our baseline estimate and performed a sensitivity analysis for the case study with 0.01 $m^2/s$, which is approximately 6 times faster than the baseline, as well as a diffusion coefficient value that is 6 times smaller than the baseline (S9 and S10 Fig). As mentioned previously, since the diffusion coefficient is directly related to the time spent by the particles in a grid cell in the air, the sensitivity analysis of diffusion coefficients is performed with the combined effects of changing aerosol decay rate (S9 Fig) and droplet deposition rate (S10 Fig) values.

**Inhalation (pickup) rate of virus from the air** ($\rho$) is defined as the ratio of human tidal volume over the cell volume per time step (here per hour). The tidal volume is defined as the amount of air that moves in or out of the lungs in each respiratory cycle and is estimated to be 0.4 L on average for an adult (Hallett, Toro, and Ashurst 2020). The respiratory rate per minute is, on average, 12 times (Hallett, Toro, and Ashurst 2020). Thus, the human ventilation volume is calculated as 0.4*12 = 4.8L per minute or 288L per hour. Individuals are thus assumed to inhale 2.304 times the cell volume (125L) each hour. The human ventilation volume increases 1.5 times while singing (Bernardi et al. 2017); hence $\rho$ can be set to 432L/125L per hour if an individual is to sing continuously for one hour.

**Proportion of pathogen secreted to hands** ($\eta$) represents the proportion of pathogen contamination that is deposited on infectious agents' hands as a result of virus emission. It is quantified as 0.15 (Kraay et al. 2018; Shuai Li et al. 2021)).

**Ratio of finger pad size over the reachable surface area** ($\pi$), **transfer efficiency between hands and surfaces** ($\theta$), and **surface touching frequency** ($\gamma$) constitute the parameter set defining the virus transfer rate between hands and surfaces. Hands can constitute an important route for SARS-CoV-2 transmission (Lin et al. 2022) and viruses on infectious people's hands can be transferred to surfaces (Winther et al. 2007).

The ratio of finger pad size over the reachable surface area ($\pi$) is calculated as 0.0196, which is the result of the calculation (average hand area*fraction of hand area used for transfer)/reachable surface area. The average hand size is set to 0.0049 $m^2$ (U.S. Environmental Protection Agency 2011; Beamer et al. 2015; Wilson et al. 2021), the fraction of hand area used for transfer is set to 0.1 (AuYeung, Canales, and Leckie 2008), and the reachable surface area is calculated as 0.5*0.5 = 0.25 metre squares based on the reachable distance assumption of 0.5 metres.

The transfer efficiency between hands and surfaces ($\theta$) depends on the surface materials. No empirical data is available on the transfer efficiency for SARS-CoV-2, but it is assumed to be diverse (Marzoli et al. 2021; Belluco et al. 2021). For instance, transfer efficiency was observed to change between <0.01%-80% for bacteriophages (Rusin, Maxwell, and Gerba 2002). Julian et al. (2010) conducted an experiment on the transfer of virus between hands and glass in both directions and found the mean transfer efficiency to be 0.23 with a standard deviation of 0.22 using microphages. Some SARS-CoV-2 modelling studies (Harvey et al. 2021; Ana K. Pitol and Julian 2021) refer to Pitol et al. (2017) and use the empirical transfer efficiency data for the transfer of bacteriophage MS2 from hand to saliva, which has a mean of 0.20. The transfer efficiency is also highly dependent on virus species. (P. Liu et al. 2013) examined transfer efficiency for norovirus and found that 0.25 of hand rinses samples are positive . We applied a transfer efficiency value of 0.25 per transfer.

The frequency of touching surfaces ($\gamma$) in a restaurant is estimated as 0.25 per minute (Lei et al. 2020), which corresponds to 15 touches per hour.

Monitoring data is being collected to get a more thorough and empirical understanding of this parameter in various settings and between individuals. Zhang et al. (2021) recently examined video data related to a restaurant associated with a COVID-19 outbreak in Guangzhou. The data shows that touch frequency for closer surfaces can go up to 10^(1.41) = 25.7. Therefore, we conducted a sensitivity analysis for the combination of virus transfer parameters ($\theta \pi \gamma$) within the range of -75% and +75% (S11 Fig).

**Fractional virus transfer rate from hands to facial membranes** ($\varepsilon$) is a parameter that is hard to quantify since the relevant data is not available. When the nature of the transfer process and the available modelling studies in the literature are considered, the virus transfer rate from hands to facial membranes has mainly **four** dimensions: (i) touching surface ratio between the hands and facial membranes, (ii) the frequency of touching the face, (iii) virus transfer efficiency from hands to face and (iv) inactivation rate of virus on the skin. Due to identifiability issues of these four dimensions, the virus transfer rate from hands to facial membranes is defined as an aggregate value of these four.

The parameters (i) and (ii) are not pathogen-specific, but dependent on human behaviour and attributes. For (i) touching surface ratio between the hands and facial membranes, many recent SARS-CoV-2 modelling studies use the fraction of front partial fingers (Harvey et al. 2021; Ana K. Pitol and Julian 2021) and for the parameter values, they refer to the empirical work AuYeung et al. (2008) and the measurements in U.S. Environımantal Protection Agency (2011). These modelling studies assume that one finger is used in each transfer, and the parameters they use correspond to a mean touching surface ratio value of 0.01. To parametrize the (ii) face touching frequency, the measurements in two previous empirical studies are frequently used in the literature: 15.7 touches per hour (Nicas and Best 2008) and 23 touches per hour (Kwok, Gralton, and McLaws 2015).

For the parameters of (iii) transfer efficiency between hands and face and (iv) the inactivation rate on the skin, reliable pathogen-specific data are not available (Kraay, 2021). For (iii) transfer efficiency, some SARS-CoV-2 modelling studies (Harvey et al. 2021; Ana K. Pitol and Julian 2021) refer to Pitol et al.(2017) and use the empirical transfer efficiency data for the transfer of bacteriophage MS2 from hand to saliva, which has a mean of 0.20. Some others (e.g., Wilson et al. 2021, (King et al. 2022) refer to the transfer efficiency of phage PRD-1 in Rusin et al. (2002), which has a mean of 0.339.

For the (iv) inactivation rate on skin, some SARS-CoV-2 modelling studies (e.g., Li et al., 2021) refer to the inactivation rate values used in MERS and other coronavirus related modelling studies (S. Xiao et al. 2018; Wolff et al. 2005), which corresponds to an exponential decay rate of 0.8 $hr^{-1}$, while some other researchers (e.g., (Kraay et al. 2021) refer to the inactivation rate of influenza (Weber and Stilianakis 2008; Nicas and Jones 2009) which corresponds to 88 $hr^{-1}$. These two values differ by two orders of magnitude.

In this context, the multiplication of best estimates of different components results in a fractional transfer rate of 0%-3.5%, however, an analysis with a wider range is required since good reference data is not available yet. The base value for effective hand-to-face transfer rate is assumed to be 1% (i.e., 1% of the virus on hands effectively reaches to facial membranes on the average), and then a sensitivity analysis is conducted for 0.1%, 0.5%, 5% and 10% (S12 Fig).

Bouchnita, Anass, and Aissam Jebrane. 2020. “A Multi-Scale Model Quantifies the Impact of Limited Movement of the Population and Mandatory Wearing of Face Masks in Containing the COVID-19 Epidemic in Morocco.” *Mathematical Modelling of*

*Natural Phenomena* 15: 31.
Campanella, Mario, Serge Hoogendoorn, and Winnie Daamen. 2009. "Improving the Nomad Microscopic Walker Model." *IFAC Proceedings Volumes* 42 (15): 12–18.
———. 2014. "The Nomad Model: Theory, Developments and Applications." *Transportation Research Procedia* 2 (January): 462–67.
Chalmet, L. G., R. L. Francis, and P. B. Saunders. 1982. "Network Models for Building Evacuation." *Management Science*. https://doi.org/10.1287/mnsc.28.1.86.
Charlotte, Nathalie. 2020. "High Rate of SARS-CoV-2 Transmission Due to Choir Practice in France at the Beginning of the COVID-19 Pandemic." *Journal of Voice: Official Journal of the Voice Foundation*, December. https://doi.org/10.1016/j.jvoice.2020.11.029.
Chen, Feng, Yongxin Gao, Zijia Wang, and Yan Liu. 2020. "Analysis on Alighting and Boarding Movement Laws in Subway Using Modified Social Force Model." *Collective Dynamics* 5 (0): 307–15.
Chen, Jingmin, and Michel Bierlaire. 2015. "Probabilistic Multimodal Map Matching With Rich Smartphone Data." *Journal of Intelligent Transportation Systems*. https://doi.org/10.1080/15472450.2013.764796.
Chen, Paul Z., Niklas Bobrovitz, Zahra Premji, Marion Koopmans, David N. Fisman, and Frank X. Gu. 2021. "Heterogeneity in Transmissibility and Shedding SARS-CoV-2 via Droplets and Aerosols." *eLife* 10 (April). https://doi.org/10.7554/eLife.65774.
Cheung C. Y., and Lam William H. K. 1998. "Pedestrian Route Choices between Escalator and Stairway in MTR Stations." *Journal of Transportation Engineering / American Society of Civil Engineers* 124 (3): 277–85.
Chin, Alex W. H., Julie T. S. Chu, Mahen R. A. Perera, Kenrie P. Y. Hui, Hui-Ling Yen, Michael C. W. Chan, Malik Peiris, and Leo L. M. Poon. 2020. "Stability of SARS-CoV-2 in Different Environmental Conditions." *The Lancet. Microbe* 1 (1): e10.
CIRES. 2020. "COVID-19 Airborne Transmission Tool Available." CIRES. June 25, 2020. https://cires.colorado.edu/news/covid-19-airborne-transmission-tool-available.
Clifton, Kelly J., Patrick A. Singleton, Christopher D. Muhs, and Robert J. Schneider. 2016. "Development of Destination Choice Models for Pedestrian Travel." *Transportation Research Part A: Policy and Practice* 94 (December): 255–65.
Coleman, Kristen K., Douglas Jie Wen Tay, Kai Sen Tan, Sean Wei Xiang Ong, Than The Son, Ming Hui Koh, Yi Qing Chin, et al. 2021. "Viral Load of SARS-CoV-2 in Respiratory Aerosols Emitted by COVID-19 Patients While Breathing, Talking, and Singing." *Clinical Infectious Diseases: An Official Publication of the Infectious Diseases Society of America*, August. https://doi.org/10.1093/cid/ciab691.
Coleman, Kristen K., Douglas Jie Wen Tay, Kai Sen Tan, Sean Wei Xiang Ong, Ming Hui Koh, Yi Qing Chin, Haziq Nasir, et al. 2021. "Viral Load of Severe Acute Respiratory Syndrome Coronavirus 2 (SARS-CoV-2) in Respiratory Aerosols Emitted by Patients with Coronavirus Disease 2019 (COVID-19) While Breathing, Talking, and Singing." *Clinical Infectious Diseases: An Official Publication of the Infectious Diseases Society of America*. https://academic.oup.com/cid/advance-article-abstract/doi/10.1093/cid/ciab691/6343417.
Daamen, Winnie, Piet H. L. Bovy, Serge P. Hoogendoorn, and Aukje Van de Reijt. 2005. "Passenger Route Choice Concerning Level Changes in Railway Stations." In *Transportation Research Board Annual Meeting*, 1930:12–20.
Dabisch, Paul, Michael Schuit, Artemas Herzog, Katie Beck, Stewart Wood, Melissa Krause, David Miller, et al. 2021. "The Influence of Temperature, Humidity, and Simulated Sunlight on the Infectivity of SARS-CoV-2 in Aerosols." *Aerosol Science and Technology: The Journal of the American Association for Aerosol Research* 55 (2): 142–53.
Danalet, Antonin, and Michel Bierlaire. 2014. "A Path Choice Approach to Activity Modeling with a Pedestrian Case Study." In *14th Swiss Transport Research Conference*. https://infoscience.epfl.ch/record/199478.
Danalet, Antonin, Bilal Farooq, and Michel Bierlaire. 2014. "A Bayesian Approach to Detect Pedestrian Destination-Sequences from WiFi Signatures." *Transportation Research Part C: Emerging Technologies* 44 (July): 146–70.
Darby, Steven, Krishnakumar Chulliyallipalil, Milosz Przyjalgowski, Paddy McGowan, Simon Jeffers, Alan Giltinan, Liam Lewis, Niall Smith, and Roy D. Sleator. 2021. "COVID-19: Mask Efficacy Is Dependent on Both Fabric and Fit." *Future Microbiology* 16 (January): 5–11.
Dias, Charitha, Muhammad Abdullah, Majid Sarvi, Ruggiero Lovreglio, and Wael Alhajyaseen. 2019. "Modeling and Simulation of Pedestrian Movement Planning Around Corners." *Sustainability: Science Practice and Policy* 11 (19): 5501.
Dijkstra, J., and A. J. Jessurun. 2016. "Pedestrian Activity Simulation in Shopping Environments: An Irregular Network Approach." In *SIMUL2016, The Eighth International Conference on Advances in System Simulation, 21-25 August 2016, Roma, Italy*, 33–39. IARIA.
Doremalen, Neeltje van, Trenton Bushmaker, Dylan H. Morris, Myndi G. Holbrook, Amandine Gamble, Brandi N. Williamson, Azaibi Tamin, et al. 2020. "Aerosol and Surface Stability of SARS-CoV-2 as Compared with SARS-CoV-1." *The New England Journal of Medicine* 382 (16): 1564–67.
Drewnick, Frank, Julia Pikmann, Friederike Fachinger, Lasse Moormann, Fiona Sprang, and Stephan Borrmann. 2021. "Aerosol Filtration Efficiency of Household Materials for Homemade Face Masks: Influence of Material Properties, Particle Size, Particle Electrical Charge, Face Velocity, and Leaks." *Aerosol Science and Technology*. https://doi.org/10.1080/02786826.2020.1817846.
Duives, Dorine C., Winnie Daamen, and Serge P. Hoogendoorn. 2013. "State-of-the-Art Crowd Motion Simulation Models." *Transportation Research Part C: Emerging Technologies* 37 (December): 193–209.
Evans, Matthew. 2020. "Avoiding COVID-19: Aerosol Guidelines." *arXiv [physics.soc-Ph]*. arXiv. http://arxiv.org/abs/2005.10988.
Fang, Zhiming, Zhongyi Huang, Xiaolian Li, Jun Zhang, Wei Lv, Lei Zhuang, Xingpeng Xu, and Nan Huang. 2020. "How Many

Infections of COVID-19 There Will Be in the 'Diamond Princess'-Predicted by a Virus Transmission Model Based on the Simulation of Crowd Flow." *arXiv [physics.soc-Ph]*. arXiv. http://arxiv.org/abs/2002.10616.

Gao, Caroline X., Yuguo Li, Jianjian Wei, Sue Cotton, Matthew Hamilton, Lei Wang, and Benjamin J. Cowling. 2021. "Multi-Route Respiratory Infection: When a Transmission Route May Dominate." *The Science of the Total Environment* 752 (January): 141856.

Gregson, Florence K. A., Natalie A. Watson, Christopher M. Orton, Allen E. Haddrell, Lauren P. McCarthy, Thomas J. R. Finnie, Nick Gent, et al. 2021. "Comparing Aerosol Concentrations and Particle Size Distributions Generated by Singing, Speaking and Breathing." *Aerosol Science and Technology: The Journal of the American Association for Aerosol Research* 55 (6): 681–91.

Guo, Zhan, and Becky P. Y. Loo. 2013. "Pedestrian Environment and Route Choice: Evidence from New York City and Hong Kong." *Journal of Transport Geography* 28 (April): 124–36.

Hallett, Sasha, Fadi Toro, and John V. Ashurst. 2020. "Physiology, Tidal Volume." In *StatPearls*. Treasure Island (FL): StatPearls Publishing.

Hamner, Lea, Polly Dubbel, Ian Capron, Andy Ross, Amber Jordan, Jaxon Lee, Joanne Lynn, et al. 2020. "High SARS-CoV-2 Attack Rate Following Exposure at a Choir Practice - Skagit County, Washington, March 2020." *MMWR. Morbidity and Mortality Weekly Report* 69 (19): 606–10.

Hänseler, Flurin S., William H. K. Lam, Michel Bierlaire, Gael Lederrey, and Marija Nikolić. 2017. "A Dynamic Network Loading Model for Anisotropic and Congested Pedestrian Flows." *Transportation Research Part B: Methodological* 95 (January): 149–68.

Harbourt, David E., Andrew D. Haddow, Ashley E. Piper, Holly Bloomfield, Brian J. Kearney, David Fetterer, Kathleen Gibson, and Timothy Minogue. 2020. "Modeling the Stability of Severe Acute Respiratory Syndrome Coronavirus 2 (SARS-CoV-2) on Skin, Currency, and Clothing." *PLoS Neglected Tropical Diseases* 14 (11): e0008831.

Harvey, Abigail P., Erica R. Fuhrmeister, Molly Cantrell, Ana K. Pitol, Jenna M. Swarthout, Julie E. Powers, Maya L. Nadimpalli, Timothy R. Julian, and Amy J. Pickering. 2021. "Longitudinal Monitoring of SARS-CoV-2 RNA on High-Touch Surfaces in a Community Setting." *Environmental Science & Technology Letters* 8,2: 168–75.

Harweg, Thomas, Daniel Bachmann, and Frank Weichert. 2021. "Agent-Based Simulation of Pedestrian Dynamics for Exposure Time Estimation in Epidemic Risk Assessment." *Zeitschrift Fur Gesundheitswissenschaften = Journal of Public Health*, April, 1–8.

Helbing, D., and P. Molnár. 1995. "Social Force Model for Pedestrian Dynamics." *Physical Review. E, Statistical Physics, Plasmas, Fluids, and Related Interdisciplinary Topics* 51 (5): 4282–86.

Hill, Michael R. 1984. *Spatial Structure and Decisionmaking Aspects of Pedestrian Route Selection through an Urban Environment*. University Microfilms.

Hinds, William C. 1999. *Aerosol Technology: Properties, Behavior, and Measurement of Airborne Particles*. John Wiley & Sons.

Hoogendoorn, Serge P., and Piet H. L. Bovy. 2005. "Pedestrian Travel Behavior Modeling." *Networks and Spatial Economics* 5 (2): 193–216.

Hoogendoorn, Serge P., Femke L. M. van Wageningen-Kessels, Winnie Daamen, and Dorine C. Duives. 2014. "Continuum Modelling of Pedestrian Flows: From Microscopic Principles to Self-Organised Macroscopic Phenomena." *Physica A: Statistical Mechanics and Its Applications*. https://doi.org/10.1016/j.physa.2014.07.050.

Hoogendoorn, S. P., and P. H. L. Bovy. 2004. "Pedestrian Route-Choice and Activity Scheduling Theory and Models." *Transportation Research Part B: Methodological* 38 (2): 169–90.

Hughes, R. L. 2000. "The Flow of Large Crowds of Pedestrians." *Mathematics and Computers in Simulation* 53 (4): 367–70.

Hughes, Roger L. 2002. "A Continuum Theory for the Flow of Pedestrians." *Transportation Research Part B: Methodological* 36 (6): 507–35.

Ijaz, M. Khalid, Raymond W. Nims, Sifang Steve Zhou, Kelly Whitehead, Vanita Srinivasan, Tanya Kapes, Semhar Fanuel, et al. 2021. "Microbicidal Actives with Virucidal Efficacy against SARS-CoV-2 and Other Beta- and Alpha-Coronaviruses and Implications for Future Emerging Coronaviruses and Other Enveloped Viruses." *Scientific Reports* 11 (1): 5626.

Jones, Benjamin, Patrick Sharpe, Christopher Iddon, E. Abigail Hathway, Catherine J. Noakes, and Shaun Fitzgerald. 2021. "Modelling Uncertainty in the Relative Risk of Exposure to the SARS-CoV-2 Virus by Airborne Aerosol Transmission in Well Mixed Indoor Air." *Building and Environment* 191 (March): 107617.

Julian, T. R., J. O. Leckie, and A. B. Boehm. 2010. "Virus Transfer between Fingerpads and Fomites." *Journal of Applied Microbiology* 109 (6): 1868–74.

Kalakou, S., Bierlaire, M. Moura, F. 2014. "Effects of Terminal Planning on Passenger Choices." In *14th Swiss Transport Research Conference*.

Kalakou, Sofia, and Filipe Moura. 2014. "Bridging the Gap in Planning Indoor Pedestrian Facilities." *Transport Reviews* 34 (4): 474–500.

Karamouzas, Ioannis, and Mark Overmars. 2010. "A Velocity-Based Approach for Simulating Human Collision Avoidance." In *Intelligent Virtual Agents*, 180–86. Springer Berlin Heidelberg.

Kasloff, Samantha B., Anders Leung, James E. Strong, Duane Funk, and Todd Cutts. 2021. "Stability of SARS-CoV-2 on Critical Personal Protective Equipment." *Scientific Reports* 11 (1): 984.

King, Marco-Felipe, Amanda M. Wilson, Mark H. Weir, Martín López-García, Jessica Proctor, Waseem Hiwar, Amirul Khan, et al. 2022. "Modeling Fomite-Mediated SARS-CoV-2 Exposure through Personal Protective Equipment Doffing in a Hospital Environment." *Indoor Air* 32 (1): e12938.

Kraay, Alicia N. M., Michael A. L. Hayashi, David M. Berendes, Julia S. Sobolik, Juan S. Leon, and Benjamin A. Lopman. 2021. "Risk for Fomite-Mediated Transmission of SARS-CoV-2 in Child Daycares, Schools, Nursing Homes, and Offices." *Emerging Infectious Diseases*. https://doi.org/10.3201/eid2704.203631.

Kraay, Alicia N. M., Michael A. L. Hayashi, Nancy Hernandez-Ceron, Ian H. Spicknall, Marisa C. Eisenberg, Rafael Meza, and Joseph N. S. Eisenberg. 2018. "Fomite-Mediated Transmission as a Sufficient Pathway: A Comparative Analysis across Three Viral Pathogens." *BMC Infectious Diseases* 18 (1): 540.

Kretz, Tobias, Andree Große, Stefan Hengst, Lukas Kautzsch, Andrej Pohlmann, and Peter Vortisch. 2011. "QUICKEST PATHS IN SIMULATIONS OF PEDESTRIANS." *Advances in Complex Systems* 14 (05): 733–59.

Kriegel, Martin, Anne Hartmann, Udo Buchholz, Janna Seifried, Sigrid Baumgarte, and Petra Gastmeier. 2021. "SARS-CoV-2 Aerosol Transmission Indoors: A Closer Look at Viral Load, Infectivity, the Effectiveness of Preventive Measures and a Simple Approach for Practical Recommendations." *International Journal of Environmental Research and Public Health* 19 (1): 220.

Kudryashova, O. B., N. V. Korovina, A. A. Pavlenko, V. A. Arkhipov, V. D. Gol'din, and E. V. Muravlev. 2015. "Aerosol Cloud Propagation in a Closed Space." *Journal of Engineering Physics and Thermophysics* 88 (3): 568–74.

Kudryashova, Olga B., Evgeny V. Muravlev, Aleksandra A. Antonnikova, and Sergey S. Titov. 2021. "Propagation of Viral Bioaerosols Indoors." *PloS One* 16 (1): e0244983.

Kwok, Yen Lee Angela, Jan Gralton, and Mary-Louise McLaws. 2015. "Face Touching: A Frequent Habit That Has Implications for Hand Hygiene." *American Journal of Infection Control* 43 (2): 112–14.

Lau, Zechariah, Ian M. Griffiths, Aaron English, and Katerina Kaouri. 2022. "Predicting the Spatio-Temporal Infection Risk in Indoor Spaces Using an Efficient Airborne Transmission Model." *Proceedings of the Royal Society A: Mathematical, Physical and Engineering Sciences*. https://doi.org/10.1098/rspa.2021.0383.

Lei, Hao, Shenglan Xiao, Benjamin J. Cowling, and Yuguo Li. 2020. "Hand Hygiene and Surface Cleaning Should Be Paired for Prevention of Fomite Transmission." *Indoor Air* 30 (1): 49–59.

Lelieveld, Jos, Frank Helleis, Stephan Borrmann, Yafang Cheng, Frank Drewnick, Gerald Haug, Thomas Klimach, Jean Sciare, Hang Su, and Ulrich Pöschl. 2020. "Model Calculations of Aerosol Transmission and Infection Risk of COVID-19 in Indoor Environments." *International Journal of Environmental Research and Public Health* 17 (21). https://doi.org/10.3390/ijerph17218114.

Lin, Yi-Chan, Rebecca J. Malott, Linda Ward, Linet Kiplagat, Kanti Pabbaraju, Kara Gill, Byron M. Berenger, et al. 2022. "Detection and Quantification of Infectious Severe Acute Respiratory Coronavirus-2 in Diverse Clinical and Environmental Samples." *Scientific Reports* 12 (1): 5418.

Li, Sheng, Joseph N. S. Eisenberg, Ian H. Spicknall, and James S. Koopman. 2009. "Dynamics and Control of Infections Transmitted from Person to Person through the Environment." *American Journal of Epidemiology* 170 (2): 257–65.

Li, Shuai, Yifang Xu, Jiannan Cai, Da Hu, and Qiang He. 2021. "Integrated Environment-Occupant-Pathogen Information Modeling to Assess and Communicate Room-Level Outbreak Risks of Infectious Diseases." *Building and Environment* 187 (January): 107394.

Liu, Han, Sida He, Lian Shen, and Jiarong Hong. 2021. "Simulation-Based Study of COVID-19 Outbreak Associated with Air-Conditioning in a Restaurant." *Physics of Fluids* 33 (2): 023301.

Liu, Pengbo, Blanca Escudero, Lee-Ann Jaykus, Julia Montes, Rebecca M. Goulter, Meredith Lichtenstein, Marina Fernandez, et al. 2013. "Laboratory Evidence of Norwalk Virus Contamination on the Hands of Infected Individuals." *Applied and Environmental Microbiology* 79 (24): 7875–81.

Liu, Xuan, John M. Usher, and Lesley Strawderman. 2014. "An Analysis of Activity Scheduling Behavior of Airport Travelers." *Computers & Industrial Engineering* 74 (August): 208–18.

Liu, Y., T. Li, Y. Deng, S. Liu, D. Zhang, H. Li, X. Wang, et al. 2021. "Stability of SARS-CoV-2 on Environmental Surfaces and in Human Excreta." *The Journal of Hospital Infection*.

Li, Xiangdong, Daniel Lester, Gary Rosengarten, Craig Aboltins, Milan Patel, and Ivan Cole. 2022. "A Spatiotemporally Resolved Infection Risk Model for Airborne Transmission of COVID-19 Variants in Indoor Spaces." *The Science of the Total Environment* 812 (March): 152592.

Lue, Gregory, and Eric J. Miller. 2019. "Estimating a Toronto Pedestrian Route Choice Model Using Smartphone GPS Data." *Travel Behaviour and Society*. https://doi.org/10.1016/j.tbs.2018.09.008.

Ma, Jianxin, Xiao Qi, Haoxuan Chen, Xinyue Li, Zheng Zhang, Haibin Wang, Lingli Sun, et al. 2020. "Coronavirus Disease 2019 Patients in Earlier Stages Exhaled Millions of Severe Acute Respiratory Syndrome Coronavirus 2 Per Hour." *Clinical Infectious Diseases: An Official Publication of the Infectious Diseases Society of America*, August. https://doi.org/10.1101/2020.05.31.20115154.

Marzoli, Filippo, Alessio Bortolami, Alessandra Pezzuto, Eva Mazzetto, Roberto Piro, Calogero Terregino, Francesco Bonfante, and Simone Belluco. 2021. "A Systematic Review of Human Coronaviruses Survival on Environmental Surfaces." *The Science of the Total Environment* 778 (July): 146191.

Matson, M. Jeremiah, M. Jeremiah Matson, Claude Kwe Yinda, Stephanie N. Seifert, Trenton Bushmaker, Robert J. Fischer, Neeltje van Doremalen, James O. Lloyd-Smith, and Vincent J. Munster. 2020. "Effect of Environmental Conditions on SARS-CoV-2 Stability in Human Nasal Mucus and Sputum." *Emerging Infectious Diseases*. https://doi.org/10.3201/eid2609.2267.

Mirzaie, Mahshid, Esmail Lakzian, Afrasyab Khan, Majid Ebrahimi Warkiani, Omid Mahian, and Goodarz Ahmadi. 2021. "COVID-19 Spread in a Classroom Equipped with Partition--A CFD Approach." *Journal of Hazardous Materials* 420: 126587.

Morawska, L., G. R. Johnson, Z. D. Ristovski, M. Hargreaves, K. Mengersen, S. Corbett, C. Y. H. Chao, Y. Li, and D. Katoshevski.

2009. “Size Distribution and Sites of Origin of Droplets Expelled from the Human Respiratory Tract during Expiratory Activities.” *Journal of Aerosol Science* 40 (3): 256–69.

Moussaïd, Mehdi, Dirk Helbing, and Guy Theraulaz. 2011. “How Simple Rules Determine Pedestrian Behavior and Crowd Disasters.” *Proceedings of the National Academy of Sciences of the United States of America* 108 (17): 6884–88.

Mürbe, Dirk, Martin Kriegel, Julia Lange, Lukas Schumann, Anne Hartmann, and Mario Fleischer. 2021. “Aerosol Emission of Adolescents Voices during Speaking, Singing and Shouting.” *PloS One* 16 (2): e0246819.

Nicas, Mark, and Daniel Best. 2008. “A Study Quantifying the Hand-to-Face Contact Rate and Its Potential Application to Predicting Respiratory Tract Infection.” *Journal of Occupational and Environmental Hygiene* 5 (6): 347–52.

Nicas, Mark, and Rachael M. Jones. 2009. “Relative Contributions of Four Exposure Pathways to Influenza Infection Risk.” *Risk Analysis: An Official Publication of the Society for Risk Analysis* 29 (9): 1292–1303.

Nicas, Mark, and Gang Sun. 2006. “An Integrated Model of Infection Risk in a Health-Care Environment.” *Risk Analysis: An Official Publication of the Society for Risk Analysis* 26 (4): 1085–96.

Oswin, Henry P., Allen E. Haddrell, Mara Otero-Fernandez, Jamie F. S. Mann, Tristan A. Cogan, Thomas G. Hilditch, Jianghan Tian, et al. 2022. “The Dynamics of SARS-CoV-2 Infectivity with Changes in Aerosol Microenvironment.” *Proceedings of the National Academy of Sciences of the United States of America* 119 (27): e2200109119.

Oyama, Yuki, and Eiji Hato. 2018. “Pedestrian Activity Model Based on Implicit Path Enumaration.” In *21st International Conference of Hong Kong Society for Transportation Studies: Smart Transportation, HKSTS 2016*, 331–38. Hong Kong Society for Transportation Studies Limited.

Pan, Jin, Charbel Harb, Weinan Leng, and Linsey C. Marr. 2020. “Inward and Outward Effectiveness of Cloth Masks, a Surgical Mask, and a Face Shield.” https://vtechworks.lib.vt.edu/handle/10919/100918.

Parisi, Daniel R., Marcelo Gilman, and Herman Moldovan. 2009. “A Modification of the Social Force Model Can Reproduce Experimental Data of Pedestrian Flows in Normal Conditions.” *Physica A: Statistical Mechanics and Its Applications* 388 (17): 3600–3608.

Paris, S., J. Pettré, and S. Donikian. 2007. “Pedestrian Reactive Navigation for Crowd Simulation: A Predictive Approach.” *Computer Graphics Forum: Journal of the European Association for Computer Graphics*. https://onlinelibrary.wiley.com/doi/abs/10.1111/j.1467-8659.2007.01090.x?casa_token=Jq6NAUJx3SIAAAAA:eXTNo-m-na9_Rysya4C60InIMdqJJ-1d1_jxhcy8oETVvn8Nwci7urkEPdn8lUKg_Ih9FgmLiF-fKKM.

Pastorino, Boris, Franck Touret, Magali Gilles, Xavier de Lamballerie, and Rémi N. Charrel. 2020. “Prolonged Infectivity of SARS-CoV-2 in Fomites.” *Emerging Infectious Diseases* 26 (9): 2256–57.

Pitol, A. K., Heather N. Bischel, Tamar Kohn, and Timothy R. Julian. 2017. “Virus Transfer at the Skin–Liquid Interface.” *Environmental Science & Technology* 51 (24): 14417–25.

Pitol, Ana K., and Timothy R. Julian. 2021. “Community Transmission of SARS-CoV-2 by Surfaces: Risks and Risk Reduction Strategies.” *Environmental Science & Technology Letters* 8 (3): 263–69.

Pöhlker, Mira L., Ovid O. Krüger, Jan-David Förster, Thomas Berkemeier, Wolfgang Elbert, Janine Fröhlich-Nowoisky, Ulrich Pöschl, et al. 2021. “Respiratory Aerosols and Droplets in the Transmission of Infectious Diseases.” *arXiv [physics.med-Ph]*. arXiv. http://arxiv.org/abs/2103.01188.

Popa, Alexandra, Jakob-Wendelin Genger, Michael D. Nicholson, Thomas Penz, Daniela Schmid, Stephan W. Aberle, Benedikt Agerer, et al. 2020. “Genomic Epidemiology of Superspreading Events in Austria Reveals Mutational Dynamics and Transmission Properties of SARS-CoV-2.” *Science Translational Medicine* 12 (573). https://doi.org/10.1126/scitranslmed.abe2555.

Qu, Yunchao, Ziyou Gao, Yao Xiao, and Xingang Li. 2014. “Modeling the Pedestrian’s Movement and Simulating Evacuation Dynamics on Stairs.” *Safety Science* 70 (December): 189–201.

Ranjan, Koushlesh. 2022. “Environmental Persistence of SARS-CoV-2 and Disinfection of Work Surfaces in View of Pandemic Outbreak of COVID-19.” https://www.intechopen.com/online-first/81375.

Ren, Chen, Chang Xi, Junqi Wang, Zhuangbo Feng, Fuzhan Nasiri, Shi-Jie Cao, and Fariborz Haghighat. 2021. “Mitigating COVID-19 Infection Disease Transmission in Indoor Environment Using Physical Barriers.” *Sustainable Cities and Society* 74 (November): 103175.

Riddell, Shane, Sarah Goldie, Andrew Hill, Debbie Eagles, and Trevor W. Drew. 2020. “The Effect of Temperature on Persistence of SARS-CoV-2 on Common Surfaces.” *Virology Journal* 17 (1): 145.

RIVM. 2020. “Ventilatie En COVID-19.” RIVM. August 21, 2020. https://lci.rivm.nl/ventilatie-en-covid-19.

———. 2021. “Nederlandse Aanpak En Maatregelen Tegen Het Coronavirus.” Rijksoverheid.nl. January 25, 2021. https://www.rijksoverheid.nl/onderwerpen/coronavirus-covid-19/algemene-coronaregels.

Robin, Th, G. Antonini, M. Bierlaire, and J. Cruz. 2009. “Specification, Estimation and Validation of a Pedestrian Walking Behavior Model.” *Transportation Research Part B: Methodological* 43 (1): 36–56.

Romero, Van, William D. Stone, and Julie Dyke Ford. 2020. “COVID-19 Indoor Exposure Levels: An Analysis of Foot Traffic Scenarios within an Academic Building.” *Transportation Research Interdisciplinary Perspectives* 7 (September): 100185.

Ronchi, Enrico, and Ruggiero Lovreglio. 2020. “EXPOSED: An Occupant Exposure Model for Confined Spaces to Retrofit Crowd Models during a Pandemic.” *Safety Science* 130 (October): 104834.

Rusin, P., S. Maxwell, and C. Gerba. 2002. “Comparative Surface-to-Hand and Fingertip-to-Mouth Transfer Efficiency of Gram-Positive Bacteria, Gram-Negative Bacteria, and Phage.” *Journal of Applied Microbiology* 93 (4): 585–92.

Sallah, Kankoé, Roch Giorgi, Linus Bengtsson, Xin Lu, Erik Wetter, Paul Adrien, Stanislas Rebaudet, Renaud Piarroux, and Jean

Gaudart. 2017. “Mathematical Models for Predicting Human Mobility in the Context of Infectious Disease Spread: Introducing the Impedance Model.” *International Journal of Health Geographics*. https://doi.org/10.1186/s12942-017-0115-7.

Sarmady, S., F. Haron, and A. Z. Talib. 2010. “Simulating Crowd Movements Using Fine Grid Cellular Automata.” In *2010 12th International Conference on Computer Modelling and Simulation*, 428–33.

Schadschneider, Andreas, Ansgar Kirchner, and Katsuhiro Nishinari. 2002. “CA Approach to Collective Phenomena in Pedestrian Dynamics.” In *Cellular Automata*, 239–48. Springer Berlin Heidelberg.

Schaik, Lucia van, Dorine Duives, Sascha Hoogendoorn-Lanser, Jan Willem Hoekstra, Winnie Daamen, Alexandra Gavriilidou, Panchamy Krishnakumari, Marco Rinaldi, and Serge Hoogendoorn. 2022. “Understanding Physical Distancing Compliance Behaviour Using Proximity and Survey Data: A Case Study in the Netherlands.” In *The 12th International Conference on Transport Survey Methods, Porto Novo, Portugal*.

Schijven, Jack, Lucie C. Vermeulen, Arno Swart, Adam Meijer, Erwin Duizer, and Ana Maria de Roda Husman. 2021. “Quantitative Microbial Risk Assessment for Airborne Transmission of SARS-CoV-2 via Breathing, Speaking, Singing, Coughing, and Sneezing.” *Environmental Health Perspectives* 129 (4): 47002.

Shatu, Farjana, Tan Yigitcanlar, and Jonathan Bunker. 2019. “Objective vs. Subjective Measures of Street Environments in Pedestrian Route Choice Behaviour: Discrepancy and Correlates of Non-Concordance.” *Transportation Research Part A: Policy and Practice* 126 (August): 1–23.

Shelat, Sanmay, Winnie Daamen, Bjorn Kaag, Dorine Duives, and Serge Hoogendoorn. 2020. “A Markov-Chain Activity-Based Model for Pedestrians in Office Buildings.” *Collective Dynamics* 5 (0): 423–30.

Smither, Sophie J., Lin S. Eastaugh, James S. Findlay, and Mark S. Lever. 2020. “Experimental Aerosol Survival of SARS-CoV-2 in Artificial Saliva and Tissue Culture Media at Medium and High Humidity.” *Emerging Microbes & Infections* 9 (1): 1415–17.

Song, Weiguo, Yanfei Yu, Weicheng Fan, and Heping Zhang. 2005. “A Cellular Automata Evacuation Model Considering Friction and Repulsion.” *Science in China. Series E: Engineering & Materials Science* 48 (4): 403–13.

Spicknall, Ian H., James S. Koopman, Mark Nicas, Josep M. Pujol, Sheng Li, and Joseph N. S. Eisenberg. 2010. “Informing Optimal Environmental Influenza Interventions: How the Host, Agent, and Environment Alter Dominant Routes of Transmission.” *PLoS Computational Biology* 6 (10): e1000969.

Stubenschrott, Martin, Christian Kogler, Thomas Matyus, and Stefan Seer. 2014. “A Dynamic Pedestrian Route Choice Model Validated in a High Density Subway Station.” *Transportation Research Procedia* 2 (January): 376–84.

Sze To, G. N., and C. Y. H. Chao. 2010. “Review and Comparison between the WellsâRiley and Dose-Response Approaches to Risk Assessment of Infectious Respiratory Diseases.” *Indoor Air* 20 (1): 2–16.

Ton, Danique, Jeroen van den Heuvel, Winnie Daamen, and Serge Hoogendoorn. 2015. “Route and Activity Location Choice Behaviour of Departing Passengers in Train Stations.” In *hEART (European Association for Research in Transportation) 2015 Conference. Copenhagen, Denmark*, 9–11.

Tordeux, Antoine, Gregor Lämmel, Flurin S. Hänseler, and Bernhard Steffen. 2018. “A Mesoscopic Model for Large-Scale Simulation of Pedestrian Dynamics.” *Transportation Research Part C: Emerging Technologies* 93 (August): 128–47.

Treuille, Adrien, Seth Cooper, and Zoran Popović. 2006. “Continuum Crowds.” *ACM Transactions on Graphics* 25 (3): 1160–68.

Ueki, Hiroshi, Yuri Furusawa, Kiyoko Iwatsuki-Horimoto, Masaki Imai, Hiroki Kabata, Hidekazu Nishimura, and Yoshihiro Kawaoka. 2020. “Effectiveness of Face Masks in Preventing Airborne Transmission of SARS-CoV-2.” *mSphere* 5 (5). https://doi.org/10.1128/mSphere.00637-20.

U.S. Environmental Protection Agency. 2011. “Exposure Factors Handbook 2011 Edition (final Report).” *Washington, DC: US Environmental Protection Agency*.

Usher, John M., Eric Kolstad, and Xuan Liu. 2010. “Simulation of Pedestrian Behavior in Intermodal Facilities.” *International Journal of Agent Technologies and Systems (IJATS)* 2 (3): 66–82.

Van Ginkel, J. T., and Evert Hasselaar. 2006. “Indoor Air Quality Influenced by Ventilation System Design.” In *ENHR Conference 2006: Housing in an Expanding Europe. Theory, Policy, Implementation and Participation, Ljubljana, July 2-5, 2006*. Citeseer. http://citeseerx.ist.psu.edu/viewdoc/download?doi=10.1.1.548.5862&rep=rep1&type=pdf.

Villena Gonzales, Roberto. 2022. “Analysis of the Capabilities of Pedestrian Dynamics for Reproducing Physical Distancing during COVID-19 in Bidirectional Flows.” https://repository.tudelft.nl/islandora/object/uuid:8ada8e0b-05ce-4a8f-9fa7-54b396a4dd14.

Vukadinovic, V., F. Dreier, and S. Mangold. 2011. “A Simple Framework to Simulate the Mobility and Activity of Theme Park Visitors.” In *Proceedings of the 2011 Winter Simulation Conference (WSC)*, 3248–60.

Vuorinen, Ville, Mia Aarnio, Mikko Alava, Ville Alopaeus, Nina Atanasova, Mikko Auvinen, Nallannan Balasubramanian, et al. 2020. “Modelling Aerosol Transport and Virus Exposure with Numerical Simulations in Relation to SARS-CoV-2 Transmission by Inhalation Indoors.” *Safety Science* 130 (October): 104866.

Weber, Thomas P., and Nikolaos I. Stilianakis. 2008. “Inactivation of Influenza A Viruses in the Environment and Modes of Transmission: A Critical Review.” *The Journal of Infection* 57 (5): 361–73.

Wells, William Firth, and Others. 1955. “Airborne Contagion and Air Hygiene. An Ecological Study of Droplet Infections.” *Airborne Contagion and Air Hygiene. An Ecological Study of Droplet Infections.* https://www.cabdirect.org/cabdirect/abstract/19562701497.

Wilson, Amanda M., Mark H. Weir, Sally F. Bloomfield, Elizabeth A. Scott, and Kelly A. Reynolds. 2021. “Modeling COVID-19 Infection Risks for a Single Hand-to-Fomite Scenario and Potential Risk Reductions Offered by Surface Disinfection.”

*American Journal of Infection Control* 49 (6): 846–48.

Winther, Birgit, Karen McCue, Kathleen Ashe, Joseph R. Rubino, and J. Owen Hendley. 2007. “Environmental Contamination with Rhinovirus and Transfer to Fingers of Healthy Individuals by Daily Life Activity.” *Journal of Medical Virology* 79 (10): 1606–10.

Wolff, Manfred H., Syed A. Sattar, Olusola Adegbunrin, and Jason Tetro. 2005. “Environmental Survival and Microbicide Inactivation of Coronaviruses.” In *Coronaviruses with Special Emphasis on First Insights Concerning SARS*, edited by Axel Schmidt, Olaf Weber, and Manfred H. Wolff, 201–12. Basel: Birkhäuser Basel.

Xiao, Shenglan, Yuguo Li, Minki Sung, Jianjian Wei, and Zifeng Yang. 2018. “A Study of the Probable Transmission Routes of MERS-CoV during the First Hospital Outbreak in the Republic of Korea.” *Indoor Air* 28 (1): 51–63.

Xiao, Yao, Mofeng Yang, Zheng Zhu, Hai Yang, Lei Zhang, and Sepehr Ghader. 2021. “Modeling Indoor-Level Non-Pharmaceutical Interventions during the COVID-19 Pandemic: A Pedestrian Dynamics-Based Microscopic Simulation Approach.” *Transport Policy* 109 (August): 12–23.

Xie, X., Y. Li, A. T. Y. Chwang, P. L. Ho, and W. H. Seto. 2007. “How Far Droplets Can Move in Indoor Environments--Revisiting the Wells Evaporation-Falling Curve.” *Indoor Air* 17 (3): 211–25.

Xu, Chunwen, Wenbing Liu, Xilian Luo, Xingyu Huang, and Peter V. Nielsen. 2022. “Prediction and Control of Aerosol Transmission of SARS-CoV-2 in Ventilated Context: From Source to Receptor.” *Sustainable Cities and Society* 76 (January): 103416.

Xu, Qiancheng, and Mohcine Chraibi. 2020. “On the Effectiveness of the Measures in Supermarkets for Reducing Contact among Customers during COVID-19 Period.” *Sustainability: Science Practice and Policy* 12 (22): 9385.

Yang, Fan, Amir A. Pahlavan, Simon Mendez, Manouk Abkarian, and Howard A. Stone. 2020. “Towards Improved Social Distancing Guidelines: Space and Time Dependence of Virus Transmission from Speech-Driven Aerosol Transport between Two Individuals.” *Physical Review Fluids* 5 (12): 122501.

Ying, Fabian, and Neave O’Clery. 2021. “Modelling COVID-19 Transmission in Supermarkets Using an Agent-Based Model.” *PloS One* 16 (4): e0249821.

Zhang, Nan, Xuguang Chen, Wei Jia, Tianyi Jin, Shenglan Xiao, Wenzhao Chen, Jian Hang, et al. 2021. “Evidence for Lack of Transmission by Close Contact and Surface Touch in a Restaurant Outbreak of COVID-19.” *The Journal of Infection* 83 (2): 207–16.

Zheng, Jing, Zhongyi Wang, Jiaming Li, Yidun Zhang, Lina Jiang, Yingying Fu, Yifei Jin, et al. 2022. “High Amounts of SARS-CoV-2 in Aerosols Exhaled by Patients with Omicron Variant Infection.” *The Journal of Infection*, February. https://doi.org/10.1016/j.jinf.2022.02.015.

Zuo, Yi Y., William E. Uspal, and Tao Wei. 2020. “Airborne Transmission of COVID-19: Aerosol Dispersion, Lung Deposition, and Virus-Receptor Interactions.” *ACS Nano* 14 (12): 16502–24.

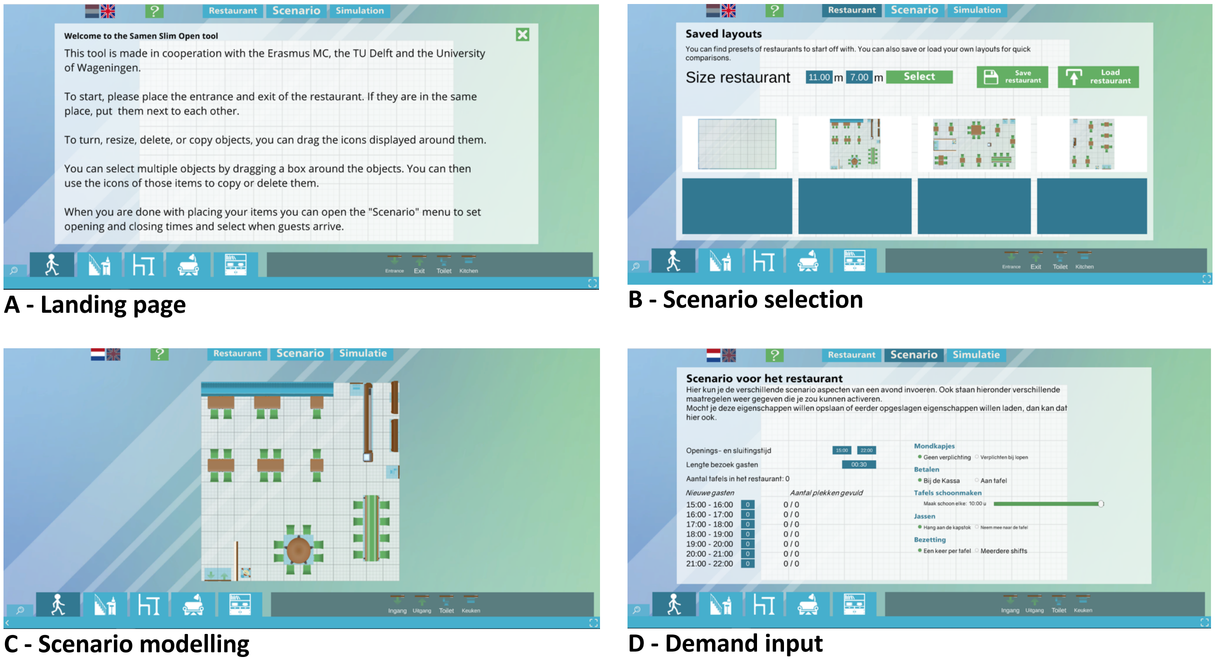


**S1 Fig. Screenshots of the SamenSlimOpen app.**
A) introduction screen, B) scene selection screen, C) scene development screen, D) developed scenario.

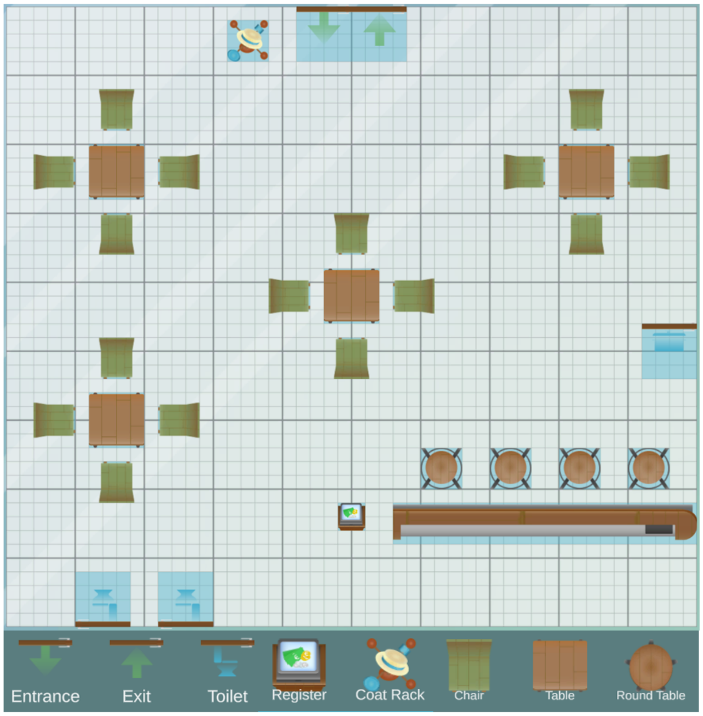


**S2 Fig. The case study restaurant layout.**
The green rectangles and round brown circles signify the seats, the green arrows the entrances, the blue toilets the entrance to the toilets and the brown rectangles the tables.

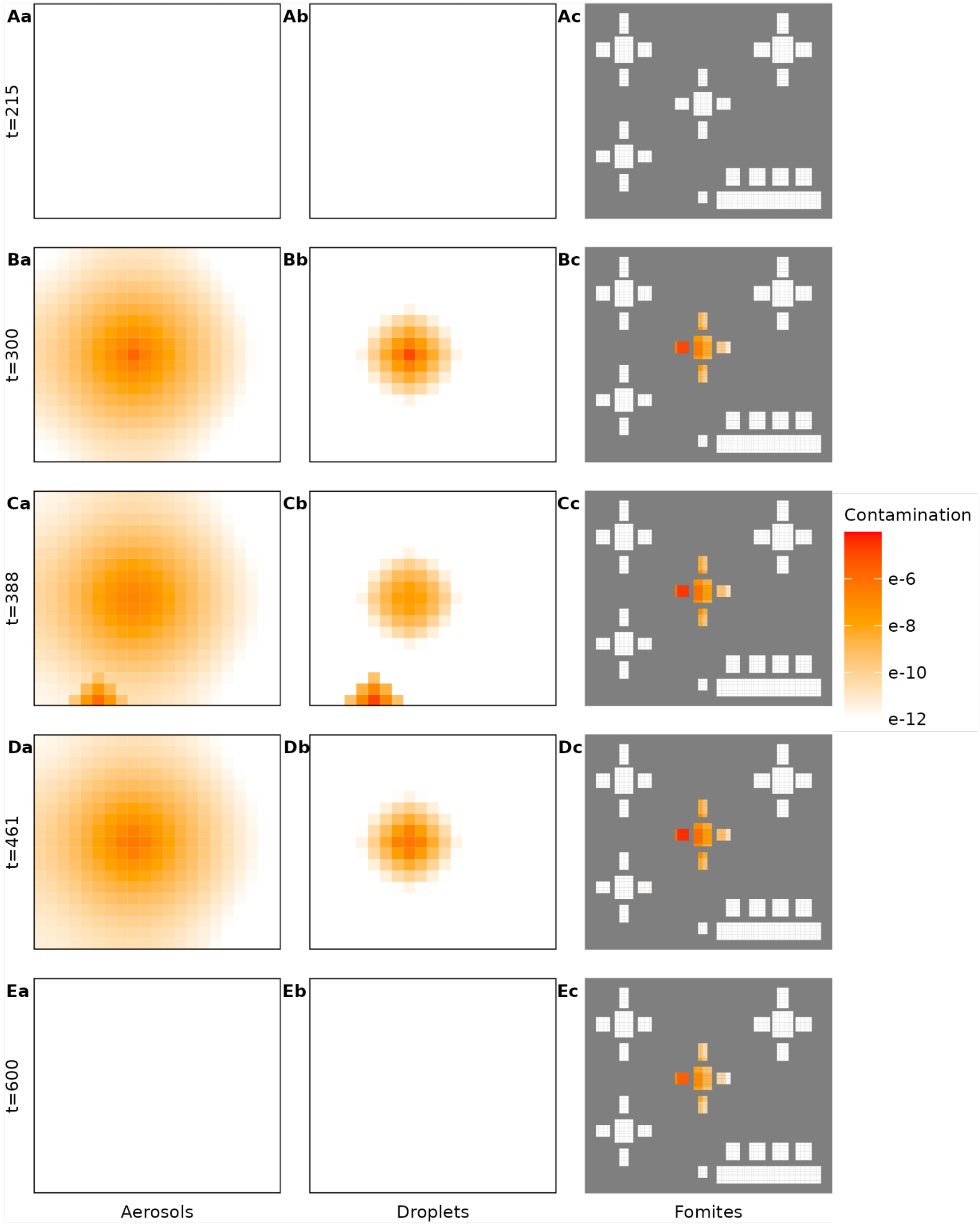


**S3 Fig. Snapshot of contamination maps in the case study.**
Virus contamination in the environment in aerosols, droplets, and on fomites over time in minutes. Contamination is expressed as the virion quantity relative to an average infectious individual's hourly emission.

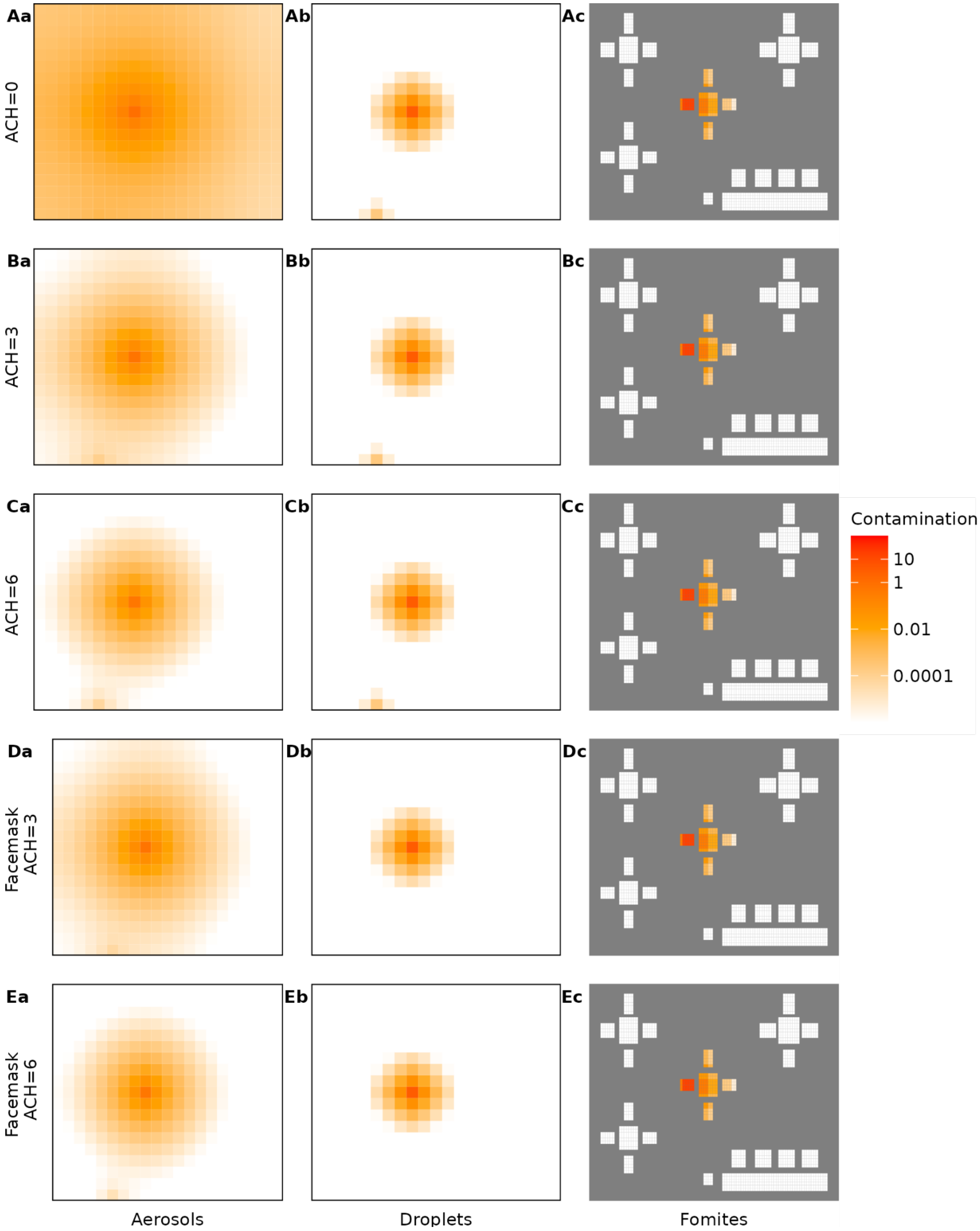


**S4 Fig. Snapshots of contamination maps in the case study for ventilation and face mask scenarios.** (A,B,C) are the scenarios where individuals do not wear face masks and ACH is 0 per hour in the restaurant in (A), 3 in (B), and 6 in (C). (D, E) are the scenarios where people wear face masks while moving and ACH is 3 per hour in the restaurant in (D) and 6 in (E). Within each scenario, the impact of intervention on viral spread is presented: (a, b, c) show virus concentration in the aerosols, droplets, and fomites, respectively.

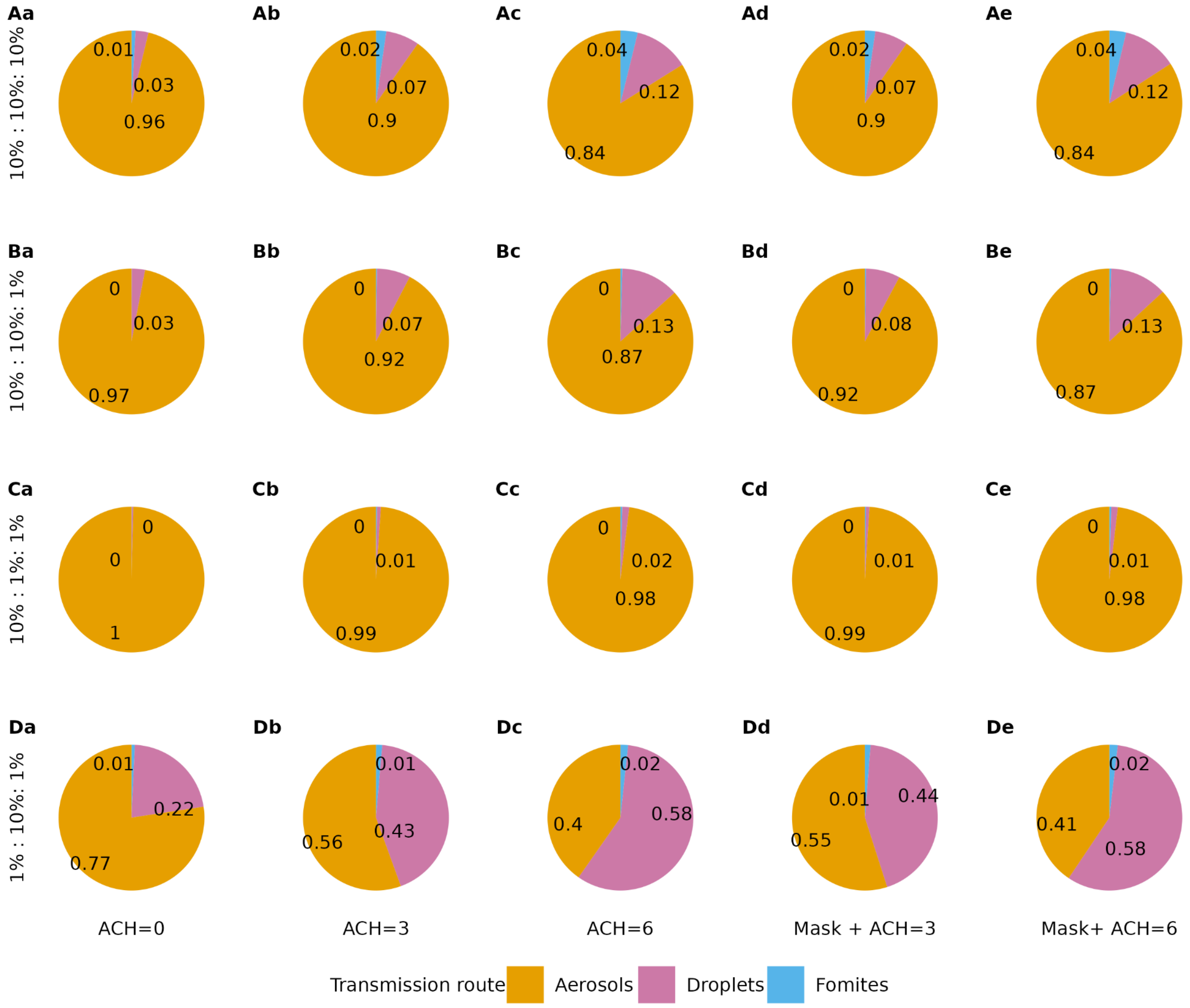


**S5 Fig. The analysis of relative contribution of transmission routes in the case study.**

Each row shows a parameters setting for $c_{route}$ (A) $c_{route}$ is the same for all routes ($c_{aerosols}$ :$c_{droplets}$ : $c_{fomites}$ is 10%:10%:10%). (B)$c_{route}$ is smaller for fomites ($c_{aerosols}$ :$c_{droplets}$ : $c_{fomites}$ is 10%:10%:1%). (C) $c_{route}$ is smaller for fomites and droplets ( $c_{aerosols}$ :$c_{droplets}$ : $c_{fomites}$ is 10%:1%:1%). (D) $c_{route}$ is smaller for fomites and aerosols ( $c_{aerosols}$ :$c_{droplets}$ : $c_{fomites}$is 1%:10%:1%). Each column shows an intervention scenario: (a) poor ventilation scenario, ACH = 0, (b) baseline scenario, ACH =3, (c) scenario with recommended ventilation, ACH =6, (d) baseline scenario with face masks worn while moving, (e) scenario with recommended ventilation and with face masks worn while moving.

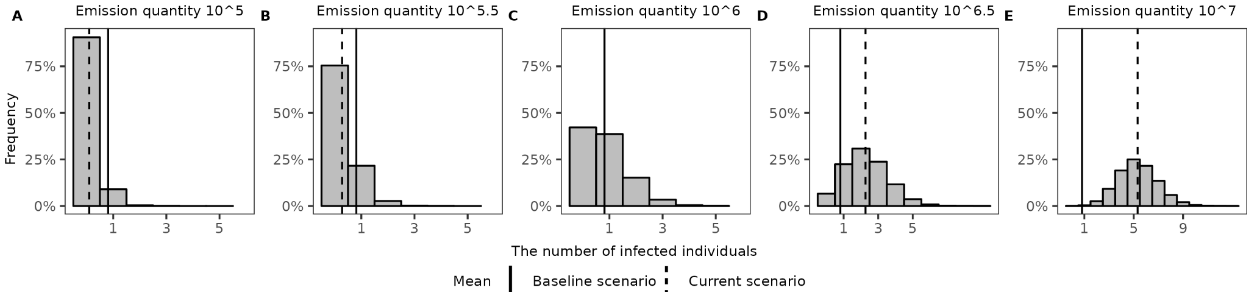


**S6 Fig. Sensitivity analysis of emission rate.**

The distributions of the expected number of infected individuals in the case study with different emission quantities $\phi$. (A) to (E) show the results for changing $\phi$ values from 10^5 to 10^7. This may reflect the heterogeneity in viral load of the index patients. The black solid lines indicate the mean value of the infected number in the baseline scenario and the dashed lines show the mean value corresponding to each respective scenario.

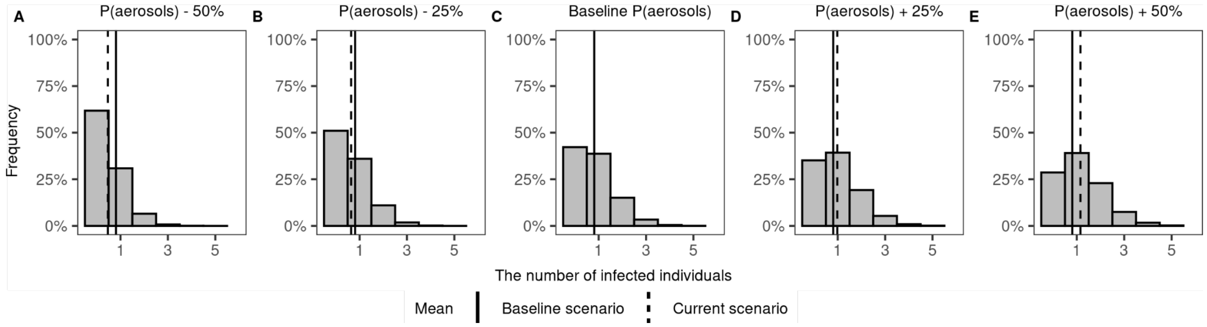


**S7 Fig. Sensitivity analysis of proportions of aerosols.**

The distributions of the expected number of infected individuals in the case study with different proportions of virus emitted in the form of aerosols . In the baseline scenario, $p_{aerosols}$ is 22.91%. (A) to (E) shows the results for from 50% lower to 50% higher (namely 11.45%, 17.18%, 22.91%, 28.63%, 34.37%) representing the heterogeneity due to respiratory activities or individual variation. The black solid lines indicate the mean value of the infected number in the baseline scenario and the dashed lines show the mean value corresponding to each respective scenario.

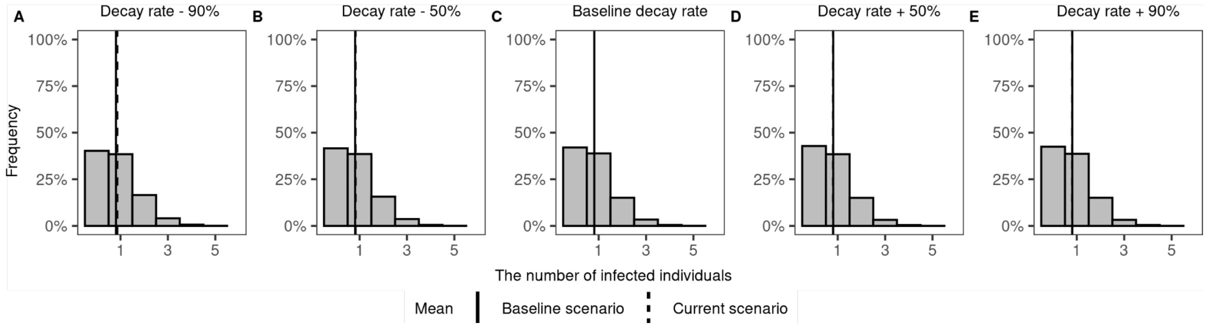


**S8 Fig. Sensitivity analysis of virus decay rate on surfaces.**

The distributions of the expected number of infected individuals in the case study with different virus decay rates on surfaces $\mu_{surfaces}$. In the baseline scenario $\mu_{surfaces}$ for wood is 0.969 per hour. (A) to (E) shows the results for changing $\mu_{surfaces}$ from 90% lower to 90% higher (namely 0.0969, 0.4845, 0.969, 1.4535, 1.8411 per hour) representing the heterogeneity due to different surface materials. The black solid lines indicate the mean value of the infected number in the baseline scenario and the dashed lines show the mean value corresponding to each respective scenario.

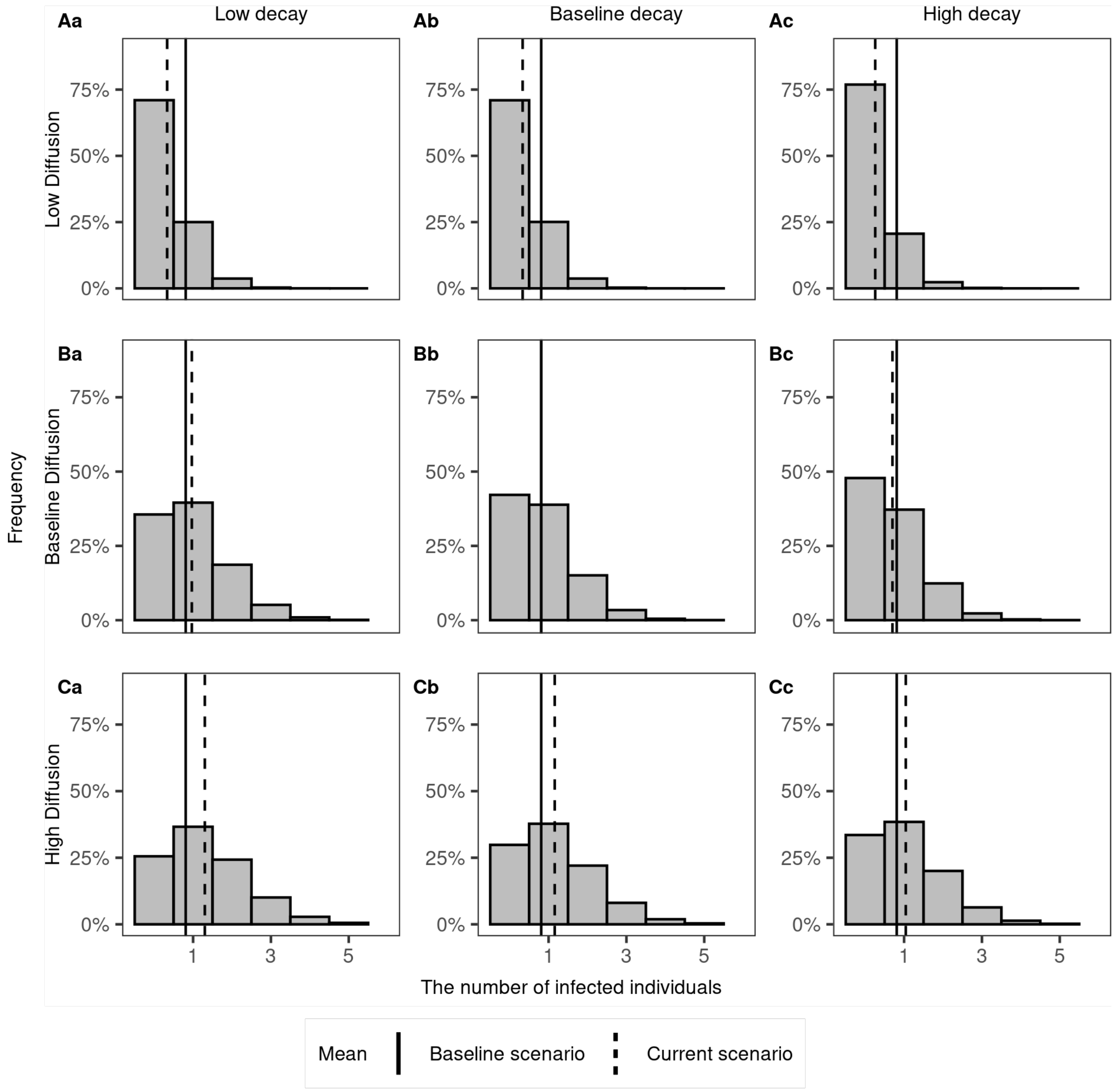


**S9 Fig. Sensitivity analysis of diffusion rate and virus decay rate in aerosols.**

The distributions of the expected number of infected individuals in the case study with different diffusion rates $D$ and virus decay rates in aerosols $\mu_{aerosols}$. Each row shows a parameter setting for diffusion: (A) Diffusion rate is 0.000278 $m^2$/s, 6 times smaller than the baseline scenario. (B) Diffusion rate is at the baseline scenario 0.0016$m^2$/s. (C) Diffusion rate is 0.01 $m^2$/s as an upper bound from literature (Olga B. Kudryashova et al. 2021), 6 times larger than the baseline scenario. Each column shows a parameter setting for virus decay rate in aerosols $\mu_{aerosols}$. (a) Decay rate is 0.755/hour, 50% lower than the baseline scenario (b) Decay rate is 1.51/hour as the baseline scenario. (c) Decay rate is 2.27/hour, 50% higher than the baseline scenario. The black solid lines indicate the mean value of the infected number in the baseline scenario and the dashed lines show the mean value corresponding to each respective scenario.

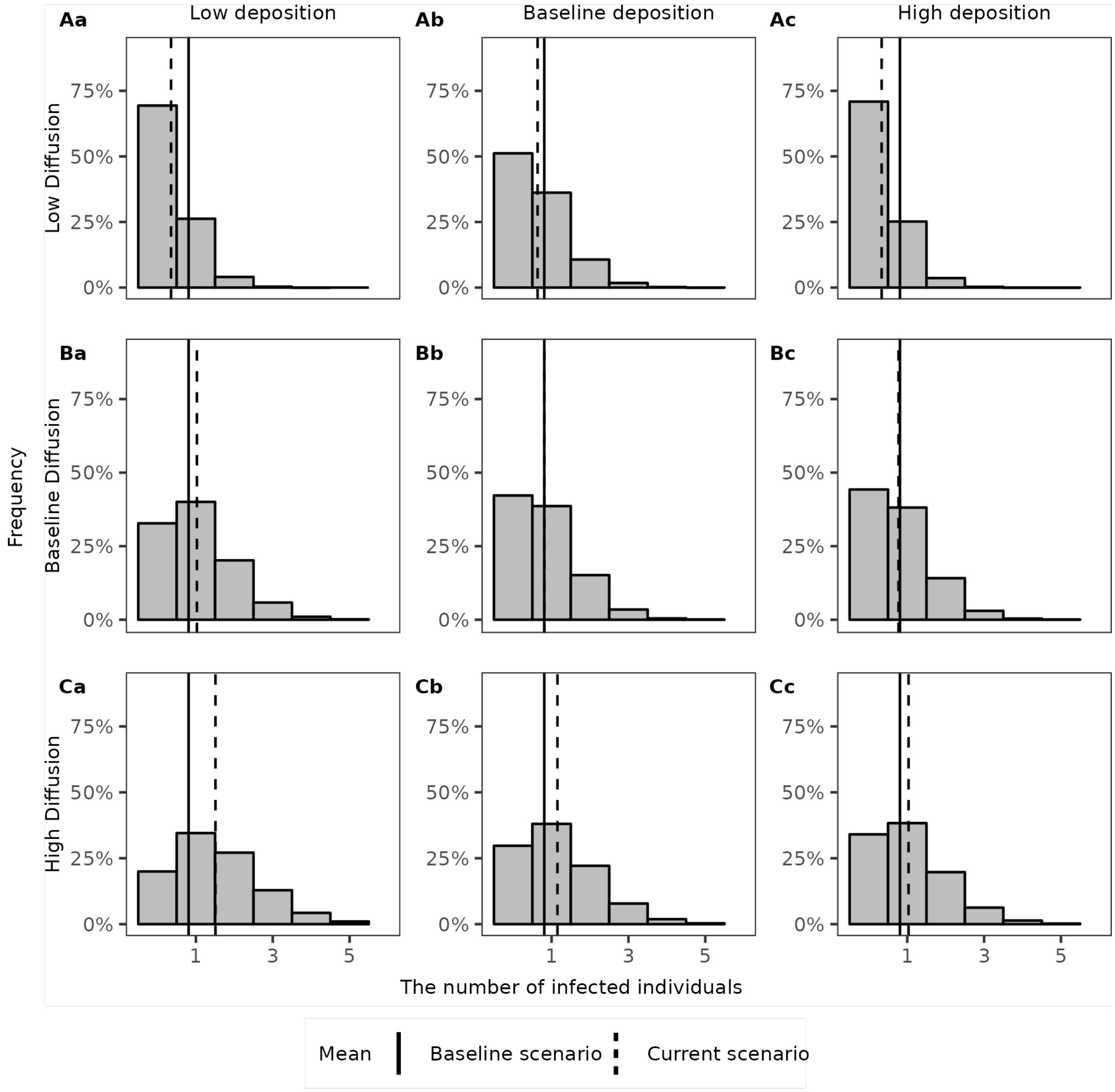


**S10 Fig. Sensitivity analysis of diffusion rate and deposition rate for droplets.**

The distributions of the expected number of infected individuals in the case study with different diffusion rates $D$ and deposition rates $\mu_{droplets}$. Each row shows a parameter setting for diffusion: (A) Diffusion rate is 0.000278 $m^2/s$, 6 times smaller than the baseline scenario. (B) Diffusion rate is at the baseline scenario 0.0016$m^2/s$. (C) Diffusion rate is 0.01 $m^2/s$ as an upper bound from literature (Olga B. Kudryashova et al. 2021), 6 times larger than the baseline scenario. Each column shows a parameter setting for deposition: (a) Deposition rate is 18.97/hour, 50% lower than the baseline scenario (b) Deposition rate is 37.93/hour as baseline scenario. (c) Deposition rate is 56.90/hour, 50% higher than the baseline scenario. The black solid lines indicate the mean value of the infected number in the baseline scenario and the dashed lines show the mean value corresponding to each respective scenario.

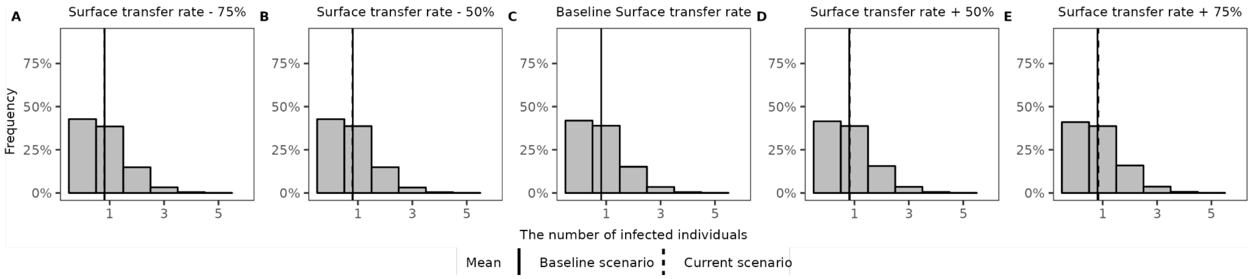


**S11 Fig. Sensitivity analysis of virus transfer rate between hand and surface.**

The distributions of the expected number of infected individuals in the case study with different virus transfer rates between hand and surface ($\theta\pi\gamma$). The baseline transfer rate between hand and surface is 0.0735 (0.0196*0.25*15) per hour. (A) to (E) shows the results for changing transfer rates from 75% lower to 75% higher (namely 0.0184, 0.0368, 0.0735, 0.1103, 0.1286 per hour) representing the heterogeneity of touching surface behaviour. The black solid lines indicate the mean value of the infected number in the baseline scenario and the dashed lines show the mean value corresponding to each respective scenario.

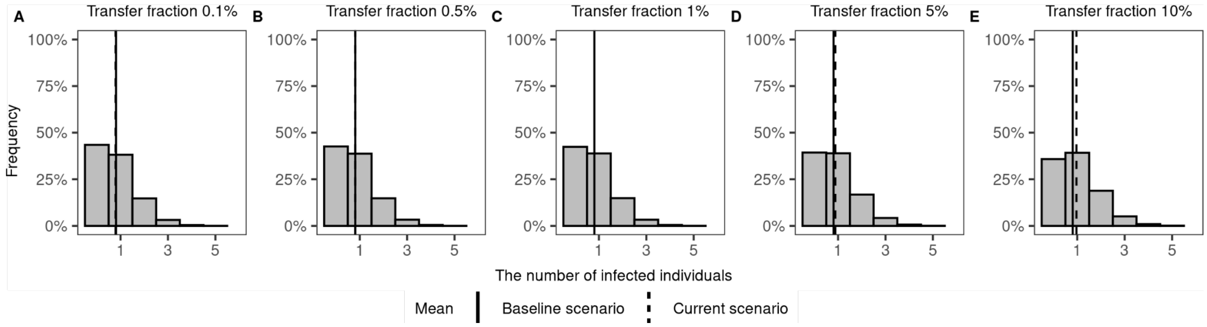


**S12 Fig. Sensitivity analysis of fractional virus transfer rate from hand to facial membranes.**

The distributions of the expected number of infected individuals in the case study with different fractional virus transfer rates from hand to facial membranes $\varepsilon$. The baseline transfer rate from hand to facial membranes is 1%. (A) to (E) shows the results for changing $\varepsilon$ from 0.1%, 0.5%, 1%, 5% and 10% representing the heterogeneity of touch face behaviour. The black solid lines indicate the mean value of the infected number in the baseline scenario and the dashed lines show the mean value corresponding to each respective scenario.
